# Urinary tract infections in children: building a causal model-based decision support tool for diagnosis with domain knowledge and prospective data

**DOI:** 10.1101/2022.04.18.22273959

**Authors:** Jessica A. Ramsay, Steven Mascaro, Anita J. Campbell, David Foley, Ariel O. Mace, Paul Ingram, Meredith L. Borland, Christopher Blyth, Nicholas G. Larkins, Tim Robertson, Phoebe Williams, Tom Snelling, Yue Wu

**Affiliations:** Wesfarmers Centre of Vaccines and Infectious Diseases, Telethon Kids Institute, University of Western Australia, Nedlands WA 6009, AU; Bayesian Intelligence Pty Ltd, Upwey VIC 3158, AU; Faculty of Information Technology, Monash University, Clayton VIC 3168, AU; Department of Infectious Diseases, Perth Children’s Hospital, Nedlands WA 6009, AU; Department of Microbiology, PathWest Laboratory Medicine WA, QEII Medical Centre, Nedlands WA 6009, AU; Department of General Paediatrics, Perth Children’s Hospital, Nedlands WA 6009, AU; School of Pathology and Laboratory Medicine, University of Western Australia, Nedlands WA 6009, AU; Emergency Department, Perth Children’s Hospital, Nedlands WA 6009, AU; Divisions of Emergency Medicine and Paediatrics, School of Medicine, University of Western Australia, Nedlands WA 6009, AU; Faculty of Health and Medical Sciences, University of Western Australia, AU; Department of Nephrology, Perth Children’s Hospital, Nedlands WA 6009, AU; Child and Adolescent Health Service, Perth Children’s Hospital, Nedlands WA 6009, AU; Sydney School of Public Health, Faculty of Medicine and Health, University of Sydney, Camperdown NSW 2006, AU; Sydney Children’s Hospital Network, NSW 2031, AU; School of Women’s and Children’s Health, The University of New South Wales, NSW 2052, AU; School of Public Health, Curtin University, Bentley WA 6102, AU; Menzies School of Health Research, Charles Darwin University, Darwin NT 0815, AU

**Author notes:** **Abbreviations:** UTI, ED, DAG, BN, CPT.

**Keywords:** DAG, causal model, Bayesian network, clinical decision support, urinary tract infection

## Abstract

**Background:** Diagnosing urinary tract infections (UTIs) in children in the emergency department (ED) is challenging due to the variable clinical presentations and difficulties in obtaining a urine sample free from contamination. Clinicians need to weigh a range of observations to make timely diagnostic and management decisions, a difficult task to achieve without support due to the complex interactions among relevant factors. Directed acyclic graphs (DAG) and causal Bayesian networks (BN) offer a way to explicitly outline the underlying disease, contamination and diagnostic processes, and to further make quantitative inference on the event of interest thus serving as a tool for decision support.

**Methods:** We prospectively collected data on children present to ED with suspected UTIs. Through knowledge elicitation workshops and one-on-one meetings, a DAG was co-developed with domain experts (the Expert DAG) to describe the causal relationships among variables relevant to paediatric UTIs. The Expert DAG was combined with prospective data and further domain knowledge to inform the development of an application-oriented BN (the Applied BN), designed to support the diagnosis of UTI. We assessed the performance of the Applied BN using quantitative and qualitative methods.

**Results:** We summarised patient background, clinical and laboratory characteristics of 431 episodes of suspected UTIs enrolled from May 2019 to November 2020. The Expert DAG was presented with a narrative description, elucidating how infection, specimen contamination and management pathways causally interact to form the complex picture of paediatric UTIs. Parameterised using prospective data and expert-elicited parameters, the Applied BN achieved an excellent and stable performance in predicting *E.coli* culture results, with a mean AUROC of 0.86 and a mean log loss of 0.48 based on 10-fold cross-validation. The BN predictions were reviewed via a validation workshop, and we illustrate how they can be presented for decision support using three hypothetical clinical scenarios.

**Conclusion:** Causal BNs created from both expert knowledge and data can integrate case-specific information to provide individual decision support during the diagnosis of paediatric UTIs in ED. The model aids the interpretation of culture results and the diagnosis of UTIs, promising the prospect of improved patient care and judicious use of antibiotics.

## 1. Introduction

Urinary tract infections (UTIs) are a common reason for children to present to hospital emergency departments (EDs) (1,2). Diagnoses of UTIs in children are made difficult because signs and symptoms are often non-specific and poorly sensitive, especially in those too young to communicate verbally. Although urine culture is considered the gold standard for the diagnosis of UTIs, urine testing may be affected by false positive and false negative results. Sample collection is challenging and urine contamination in children is frequent and can cause false positive diagnoses or obscure true positive infections, resulting in inappropriate treatment (4). Urine testing may also be affected by false negatives due to prior use of antibiotics and low bacterial counts (3). Management of UTIs in children requires timely decisions that balance the risks of secondary bacteraemia and sepsis if appropriate treatment is delayed, and the potential side effects of those treatments, as well as the growing public health risks from antimicrobial resistance associated with indiscriminate treatment (3).

Formulating a diagnosis relies on gathering, requesting, and synthesising information from multiple sources under time and resource constraints. Cognitive heuristics (i.e. short cuts) allow decisions to be made quickly and with little information and high uncertainty; however, these heuristics may be biased and are thought to contribute to 75% of misdiagnoses (4,5). The management of children with suspected UTI in the ED could benefit from decision support based on quantitative modelling. A number of predictive models have been constructed to aid the diagnosis and management of UTIs in children, with varying success. Individual biomarkers have been proposed for guiding diagnosis (6,7), treatment and prognostication, while others propose combining routinely collected information to provide quantitative risk-based assessments (8,9). The lack of explanability and user engagement may be the reason many predictive models, regardless of their accuracy, fail to be successfully implemented or utilised (10–13). Causal directed acyclic graphs (DAGs) can be used to map and describe effects centred around a causal question of interest (14), providing a potential way to address the lack of explanability.

Causal DAGs are a graphical representation of variables of interest and their relationships with each other, depicted by a series of nodes (variables) and arrows (the causal relationships between the connected variables) (15). They assist in understanding when and how observing one variable should change our expectation of another, either because the first variable *causes* the second, the first is *caused by* the second, each is *caused by a third* variable, or because each *shares an effect* which is also observed. Bayesian network (BN) models extend DAGs by quantifying the strength and direction of the cause-effect relationships between variables using conditional probability tables (CPTs), capturing a deeper understanding of a problem (16,17). When the relationships between all relevant observable and unobservable (latent) variables are organised under a causal BN framework, observed variables (data) can then be used to make probabilistic inference about missing variables that are either unobservable and must always be inferred (e.g., latent states), or those that are potentially observable but not yet observed (e.g., future outcomes). They provide an approach for designing decision support tools by predicting unobserved variables using available data. Causal BN models can be synthesised to describe a complex problem by combining expert opinion on the qualitative structure (i.e. the DAG) with expert opinion and/or data to parameterize it (17). This approach compensates for data limitations to improve model predictions and may help to illuminate the clinical problem at hand and increase the likelihood that any decision support tools arising from these models will be used in clinical care.

Causal BN models thereby offer a way of organising information in a coherent way that captures the complex relationships amongst variables relevant to the problem domain of paediatric UTIs. Here we describe the methodological process of building a causal BN based decision support tool for diagnosing the causative pathogens for children who present to ED for suspected UTI. We illustrate how an expert-elicited causal DAG can be translated into an applied BN model parameterised with a prospective paediatric cohort. We discuss the potential use of the applied BN model in clinical settings with the aim of guiding the diagnosis and management of UTI in children.

## 2. Methods

This project is described in three phases to illustrate how prospective cohort data (2.1) and an expert-elicited causal DAG (2.2) can be utilised to derive a clinical decision support BN quantifying the strength of these relationships (2.3).

### 2.1 The prospective paediatric emergency department cohort

Our prospective cohort enrolled children from the ED of Western Australia’s sole tertiary public children’s hospital (Perth Children’s Hospital). The study aimed to capture clinical and laboratory information about UTIs and their risk factors from paediatric ED clinicians, laboratory results, and from parents of children with a suspected UTI. A child was included if they were aged less than 13 years, presented to the ED with a suspected UTI, had urine collected for laboratory culture and susceptibility testing, were prescribed empiric antibiotics for their suspected UTI, and had informed consent provided by their legal guardian. Participants could be re-enrolled if they presented to the ED at least 14 days after their initial presentation. Ethics approval was granted by the Child and Adolescent Health Service Human Research Ethics Committee (EC00268).

Electronic and paper medical records were systematically reviewed to capture the participant’s clinical history including their demographics, reported signs and symptoms, clinical observations, laboratory results, and treatments prescribed. Parents were surveyed at enrolment to identify any additional risks factors for antimicrobial resistance and 14 days after presenting to the ED to ascertain treatment outcomes. Samples were processed, analysed and reported by the local laboratory per their standard procedures. Additional file 1 provided a detailed schematic of the participants enrolment and data collection.

### 2.2 Qualitative model: the Expert DAG

A qualitative causal DAG was constructed based on knowledge elicited from local clinical experts over multiple iterations (the ***Expert DAG***). The experts were chosen to represent a range of health professionals typically involved in the diagnosis and management of children with UTIs at a tertiary hospital, and therefore the intended end-users of a decision support tool resulting from this work. The experts were from paediatric emergency medicine, microbiology and infectious diseases, general paediatrics, nephrology, epidemiology and medical laboratory science.

The elicitation rested on an initial causal framework based on preliminary insights from the prospective cohort data and mixed domain and modelling knowledge from a core team (YW, JAR, SM, TS). Proposed relationships from this initial framework were then confirmed, corrected, or expanded after input from the broader expert group (DF, AJC, PI, MLB, CB, NGL, TR, AOM, PW). Many causal relationships between model variables were fairly intuitive and not controversial, meaning the relationships were clear (often visible) events occurring in clear temporal sequence. Therefore, elicitation of the model structure occurred with moderated discussion where a full Delphi protocol was not warranted. Additionally, discussions within a diverse expert group allowed consensus to be achieved, with specialty input only requested when needed, replicating decision-making processes in clinical care.

The outcome DAG elicited from the experts was then refined by the core team and re-presented in a written format, with each causal relationship depicted explicitly described. Further iteration was sought via written feedback and one-on-one expert and core team discussions. The resultant final Expert DAG describing the diagnosis and management of UTIs in children is described in Section 3.2.

### 2.3 Quantitative model: the Applied BN

The final Expert DAG was converted into an application-oriented BN (the ***Applied BN***), designed to illustrate how BN models can provide clinical decision support for the diagnosis and management of suspected UTI in children who present to the ED. Information from both the Expert DAG and the prospective cohort data were integrated to inform the selection of Applied BN variables. Conversion of the Expert DAG took into consideration: how a particular variable is relevant to the Applied BN’s purpose; how it could be matched to available data; and how it could help simplify parameterisation or computational workload. This process frequently involved simplifications by removing and merging variables, and expansions by splitting and adding variables. All changes during the conversion ensured the structure of the Applied BN was compatible with the Expert DAG, meaning all the elicited causal relationships were preserved either by explicit causal links or, where it was considered necessary, non-causal approximations.

The Applied BN was parameterised using data from the prospective cohort. In many cases, a variable’s probability conditional on its parents (predecessor node) could be estimated directly from the data. However, some of the variables in the Applied BN are latent, as they play crucial explanatory or simplification roles, and parameterisation in such cases is less straightforward. There are two kinds of latent parameters associated with latent variables: parameters that quantify the relationship of the latent variable with its parents; and parameters that quantify the relationship of the latent variable with its children (nodes extending from other nodes). In most cases, latent parameters were handled by eliciting estimated probabilities from experts and using these estimates as seeds to the expectation maximisation (EM) algorithm (18). Specifically, parameterisation surveys were created and issued to experts to elicit parameters for all latent variables and these parameters were used to inform the corresponding CPTs. In most cases, these CPTs constituted priors that were further updated by the prospective cohort data, while in other cases, the CPTs were kept fixed. In addition, one group of latent parameters were determined separately, making use of EM in the form of a clustering algorithm to “complete” the data (see Section 3.3 for description). Additional file 2 includes the full list of survey questions used to elicit parameters for the Applied BN.

The Applied BN was evaluated from the perspective of both (numerical) accuracy and clinical usefulness. BN predictions for a selected set of target variables (e.g., pathogen-specific urine culture results) were compared with the observations of those variables captured in the prospective cohort study. The difference between the BN predictions and observations were described using two metrics based on 10-fold cross-validation, namely the area under the receiver-operator curve (AUROC) and the log loss (19), both intended to measure the performance characteristics of the model, though each in different ways. A sensitivity analysis was conducted on the conditional probability parameters with a high degree of uncertainty, using variance-based sensitivity analysis (VBSA) (20,21). VBSA allows the distribution of several input parameters to be investigated simultaneously to help understand how changes influence the BN target predictions in the CPTs. The clinical experts evaluated the clinical usefulness of the BN via a validation workshop where relationships and concepts were checked and refined. Three scenarios were simulated to demonstrate how the Applied BN might be used for clinical decision support for a child presenting to the ED with a suspected UTI.

## 3. Results

### 3.1 Prospective paediatric cohort

From May 2019 to November 2020, 391 children were enrolled in the prospective cohort study. This accounted for 431 UTI episodes, where the mean age at presentation was 3.9 years old (Interquartile Range, IQR, 0.7 - 6.2) and 316 (73%) were girls. A prior history of UTI or urinary tract pathology (e.g., neuropathic bladder, phimosis, renal agenesis, dysplasia) were reported in 197 (46%) of participants according to their medical notes or reported by their parent in the study survey. Commonly reported symptoms on ED presentation included parent-reported fever (269, 62%), nausea and/or vomiting (169, 39%), poor oral intake (161, 37%), abdominal pain (144, 33%), and pain or discomfort referrable to the urinary tract (148, 34%). Symptoms varied significantly between those < 2 years old and those ≥ 2 years old (Table 1). Children were prescribed antibiotics during their episode of care as per the inclusion criteria, where broad spectrum^1^ antibiotics were prescribed in 32% of children. Among the 431 urine samples collected in the ED, 219 (51%) reported pure growth, 150 (35%) reported no growth and 7 (2%) reported mixed growth, while urine culture data was unavailable for 56 episodes (13%). *Escherichia coli* (*E.coli*) was the most common bacteria reported accounting for 204 (47%) of total episodes and 90% of positive urine samples (204/226). Other Gram negative organisms (e.g. *Proteus mirabilis, Enterobacter cloacae, Pseudomonas aeruginosa*) and Gram positive organisms (e.g. *Staphylococcus aureus, Enterococcus faecalis*) were isolated in 4% and 3% of total episodes, attributing to 7% (16/226) and 6% (13/226) of positive samples, respectively. Antibiotic use prior to ED presentation was reported in 61 (14%) of episodes and was negatively associated with urine culture (Table 1).

**Table 1.** Prospective cohort study summary statistics. Unless stated otherwise, all percentages were calculated using positively reported observations within each age group (i.e., as a percentage of the 179 cases for <2yo, and 252 cases for >=2yo). Of note, when a variable (e.g., abdominal pain) was not reported, it’s likely that the child reported no pain (confirmed negative observation) or the data was missing (e.g., not queried or recorded by the treating doctors).

| **Demographics and clinical history** |  |  |
| --- | --- | --- |
| **Subgroup by age group** | **<2yo 179 (41.5% of total 431 episodes)** | **≥2yo 252 (58.5% of total 431 episodes)** |
| Female | 101 (56%) | 215 (85%) |
| Prior urinary tract pathology (including previous UTI) | 55 (31%) | 142 (56%) |
| On antibiotics at ED presentation<sup>1</sup> |  |  |
| Broad | 6 (3%) | 14 (6%) |
| Narrow | 15 (8%) | 26 (10%) |

<sup>1</sup> The specified antibiotic was classified as narrow (<=3) or broader (>3) according to published Antibiotic Spectrum Index (22). Narrow: Amoxicilin, Trimethoprim, Benzylpenicillin, Cefalexin, Cefazolin, Erythromicin. Broad: Amoxicillin + Clavulanic acid, Trimethoprim + Sulfamethoxazole, Co-trimoxazole, Amikacin, Cefepime, Cefotaxime, Ceftazidime, Ceftriaxone, Ciprofloxacin, Colistin, Ertapenem, Gentamicin, Meropenem, Moxifloxacin, Nitrofurantoin, Norfloxacin, Piperacillin + Tazobactam, Tobramycin, Vancomycin

|  |  |  |
| --- | --- | --- |
| ***Clinical symptoms recorded*** |  |  |
| Pain or discomfort referable to the urinary tract (e.g., dysuria, genital pain) | 11 (6%) | 137 (54%) |
| Parent reported fever | 137 (77%) | 132 (52%) |
| Temperature >38°C | 43 (24%) | 64 (25%) |
| Abdominal pain | 5 (3%) | 139 (55%) |
| Foul smelling urine | 34 (19%) | 27 (11%) |
| Haematuria | 6 (3%) | 17 (7%) |
| Irritable | 72 (37%) | 19 (8%) |
| Lethargy | 52 (29%) | 51 (20%) |
| Nausea/vomiting | 76 (42%) | 93 (37%) |
| Poor oral intake | 85 (47%) | 76 (30%) |
| Diarrhoea | 25 (14%) | 12 (5%) |
| Respiratory symptoms | 46 (26%) | 43 (17%) |
| ***ED Investigations and management recorded*** |  |  |
| C-reactive protein |  |  |
| ≥15 mg/L | 49 (27%) | 29 (12%) |
| Investigation not done | 108 (60%) | 206 (82%) |
| Leucocyte count |  |  |
| ≥10 x10<sup>9</sup>/L | 57 (32%) | 30 (12%) |
| Investigation not done | 109 (61%) | 209 (83%) |
| Neutrophil count |  |  |
| ≥8 x 10<sup>9</sup>/L | 26 (15%) | 28 (11%) |
| Investigation not done | 109 (61%) | 209 (83%) |
| Broad spectrum<sup>1</sup> antibiotic empirically prescribed | 57 (32%) | 82 (33%) |
| Patients discharged after ED consult | 108 (60%) | 217 (86%) |
| ***Urine analysis*** |  |  |
| Method of urine specimen collection |  |  |
| Clean catch | 63 (35%) | 112 (44%) |
| Catheter | 70 (39%) | 16 (6%) |
| Suprapubic aspirate | 2 (1%) | 0 (0%) |
| Bacteria seen on microscopy | 110 (66%) | 94 (37%) |
| >100 leucocytes per high power field | 107 (60%) | 145 (57%) |
| Moderate epithelial cells on microscopy | 21 (12%) | 32 (13%) |
| Leucocyte esterase (3+) on urine dipstick | 51 (28%) | 100 (40%) |
| Nitrites detected on urine dipstick | 64 (36%) | 76 (30%) |
| ***Urine culture*** |  |  |
| No growth | 47 (26%) | 103 (41%) |
| *E.coli* | 97 (54%) | 107 (42%) |
| Gram-negative bacteria (other than *E.coli*) | 6 (3%) | 10 (4%) |
| Gram-positive bacteria | 5 (3%) | 8 (3%) |
| **Subgroup by antibiotics use prior to ED** | **On antibiotic 61 (14%, n=431)** | **Not on antibiotic 342 (79%, n=431)** |
| No growth | 35 (57%, n=61) | 106 (31%, n=342) |
| *E.coli* | 13 (21%, n=61) | 177 (52%, n=342) |
| Gram-negative bacteria (other than *E.coli*) | 4 (7%, n=61) | 11 (3%, n=342) |
| Gram-positive bacteria | 2 (3%, n=61) | 11 (3%, n=342) |

### 3.2 Expert DAG description

The Expert DAG comprising 29 variables represents a mechanistic causal model of UTI infection, diagnosis and management of children presenting to an ED (Figure 1). A detailed variable dictionary for the Expert DAG is provided in Additional file 3. The model can be divided into the infection, contamination, and management pathways.

**Figure 1.**
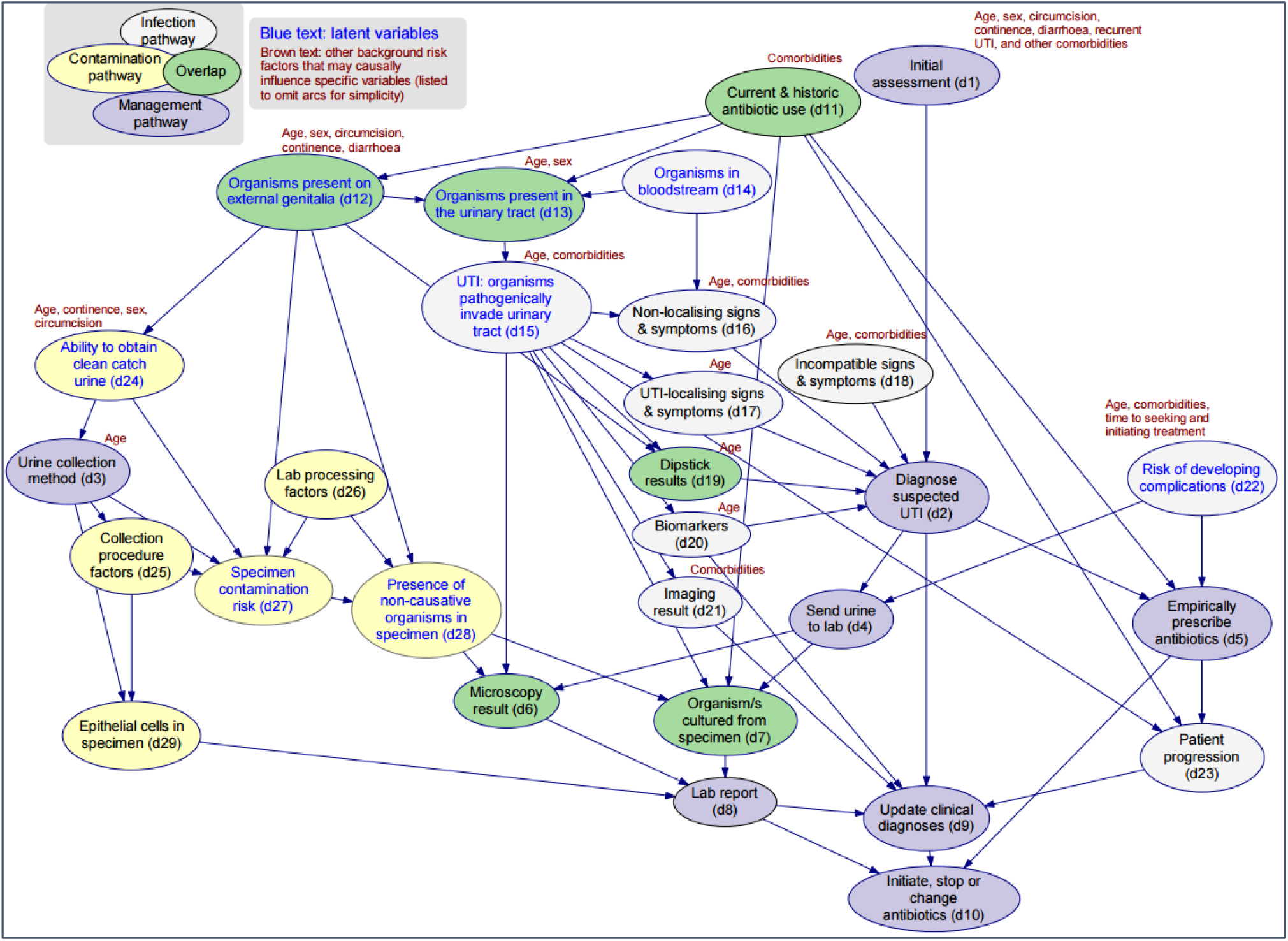
The Expert DAG v11.1. The expert-elicited causal directed acyclic graph describing the relationships between infection (white), specimen contamination (yellow) and UTI management (purple) in children, in particular, variables that fell into more than one pathways were indicated in green. Note: Numbers within the model nodes correspond with the narrative description. A detailed variable dictionary is provided with the supplementary material: Additional file 3. The source model file for the Expert DAG can be accessed via https://osf.io/8taqy/.

#### The Infection Pathway

The infection pathway describes predisposing background factors and the pathophysiology of infection, and how a UTI gives rise to signs, symptoms and laboratory evidence. For a UTI to occur, organisms must be present in the urinary tract (d13), usually from ascension of organisms from the external genitalia (d12) or, on rare occasions, from haematogenous seeding of the upper urinary tract with organisms from the bloodstream (d14) which then infect the urinary tract (d15) (23). Infection here is a ‘latent’ event, meaning that although it may be inferred from evidence with varying confidence, it generally cannot be directly observed; importantly we separate the *existence* of a *UTI* (infection pathway, d15), from the *diagnosis* of a *suspected UTI* (management pathway, d2) based on the presence or absence of various signs, symptoms, dipstick test and laboratory results. Age and UTI-relevant comorbidities (such as structural or functional abnormalities of the urinary tract) influence the probability of a UTI in a given child, due to their predisposing effect (24). Infection of the urinary tract typically provokes an inflammatory response which may manifest as UTI-localising signs and symptoms (d17) caused by inflammation of the urinary tract, and/or non-localising signs and symptoms (d16) caused by systemic inflammation. In children, especially those too young to communicate verbally, UTI-localising symptoms may be difficult to ascertain, forcing clinicians to assess observable signs and symptoms that are non-specific and non-localising such as fever and irritability, and which are shared with other conditions (25). Where incompatible signs and symptoms (d18) are present - those not typically associated with a UTI (e.g., respiratory symptoms), the diagnosis is dependent on the probability of alternative diagnoses that may provide a better explanation for the child’s presentation.

#### The Contamination Pathway

The practical definition of urinary contamination varies widely across the literature and in practice (26,27). Contamination and infection are often considered mutually exclusive, but in reality organisms cultured from urine samples may be pathogens, contaminants, or both. In our model, contamination is treated as a latent event, and describes the presence of non-causative organisms in a urine specimen (d28). Contamination usually occurs at the time of collection when organisms present superficially on the external genital area (d12) become mixed with the ‘clean’ urine sample from the bladder (in this context, ‘clean’ means the specimen is free from contaminants, not that it is free of organisms). A child’s age, sex, and for boys, circumcision status, can directly influence both the density of any organisms present on the external genitalia (d12) and the ability to produce a clean urine sample (d24). Incontinence and/or diarrhoea may increase the density of organisms present on the external genitalia (d12), increasing the risk of specimen contamination (d27), and possibly also the risk of infection of the urinary tract (d15) via the ascending route (d12).

The probability of contamination is strongly influenced by the urine collection method (d3). Within the model, the latent concept of specimen contamination risk (d27) represents all factors contributing to contamination which, if true, increases the probability of the presence of non-causative organism in the specimen (d28). Laboratory processing factors (d26) representing any process that may introduce (rare in most laboratories) or concentrate non-causative organisms in the specimen (d28) from when the urine arrives in the laboratory to its final reporting. This may include delays in sample processing or refrigeration and improper aseptic technique.

#### The Management Pathway

The existence of a UTI cannot be known with absolute certainty, and a clinician’s belief (or judgement) about its presence or absence may vary over time, perhaps related to evolving evidence. It may be suspected on the clinician’s initial assessment (d1) based on the child’s history and background risk factors. As more evidence is gained via the elicitation of symptoms and signs and from investigations, a working or provisional diagnosis of UTI is made (d2) – thus, the suspicion based on the initial assessment (d1) is updated. A urine specimen may be sent to the laboratory (d4) and if the suspicion of UTI is sufficiently high, empiric antibiotics may be prescribed (d5) even before the urine testing results are known. Management decisions are also influenced by whether the clinician believes that there is a high risk of the patient having or developing complications (d22). In the model, this is represented as a latent concept that describes the risk of progressing to severe complications. This risk is largely driven by a child’s age, the time delay to seeking and/or initiating treatment, and the presence of comorbidities such as abnormalities of the urinary tract or immune system.

Interpretation of the presence, type and density of growth cultured from a urine specimen (d7) is difficult, as this is where the contamination and infection pathways converge. Information regarding these pathways is not normally available to the laboratory scientist deciding how to report the results (d8) of the urine test. Thus, if an organism is isolated with evidence of an inflammatory response (e.g. pyuria) on microscopic analysis (d6), the probability that the cultured organism is causative is high and therefore it is reported as significant in the laboratory report (d8) and antimicrobial susceptibility results are also reported. In contrast, the isolation of multiple organisms is typically reported as a ‘mixed growth’, precluding either the confirmation or exclusion of a UTI.

A final updated clinical diagnosis (d9) is made when outstanding evidence or other information is available. The existence of a UTI directly influences the urine laboratory report (d8), any biomarker (d20) and imaging results (d21), as well as the subsequent clinical progress of the child (d23) with or without antibiotic treatment. A clinician uses these observations to further update their belief about the probability that the patient has a UTI, together with any antimicrobial susceptibility data from the laboratory report (d8) to decide whether to initiate, stop or change the antibiotic prescription (d10).

### 3.3 Applied BN for decision support

The Applied BN represents a demonstrative decision support tool using the Expert DAG that aims to help determine if a child truly has a UTI and if so, the likely causative pathogen. To develop this BN, variables in the Expert DAG were mapped to available data from the prospective cohort. Conversion of the Expert DAG into the Applied BN required simplification and expansion, whilst ensuring compatibility and preservation of the causal knowledge. Illustrative steps are summarised along the top of Figure 2. In this example, a fragment of the Expert DAG is selected (step a) that describes the presence or colonisation of bacterial pathogens on the external genitalia and in the urinary tract using two variables (d12 and d13), with an arc between them indicating that pathogens may spread from the genitalia to the urinary tract. In addition, there is depicted another possible (albeit uncommon) pathway for a pathogen to reach the urinary tract haematogenously via the bloodstream (d14). For simplicity, d14 was removed, and since this left only one explicit pathway, d12 and d13 were combined into a single variable that broadly describes local colonisation (step b). The local colonisation variable was then expanded (step c) into three nodes (b7-9) to describe local colonisation for three specific pathogen groups which are of key interest and that not only affect the probability of developing UTI, but may also constitute the causative pathogen if UTI is present (b10). Variable states were then selected (step d), typically to match the data where possible. However, in the case of latent states, this was not possible and the goal instead was to represent key divisions within each variable while minimising the demand on the latent parameterisation process. Here, each local colonisation variable is latent and has been assigned two states (High and Low), with the causative pathogen variable being assigned four states (one state for each possible causative pathogen plus a state for no pathogen/no UTI). The causative pathogen was assumed to be singular and mutually exclusive, i.e., assuming no co-infection of the urinary tract by two or more pathogens. Additional file 4 includes a full list of differences between the two models.

**Figure 2.**
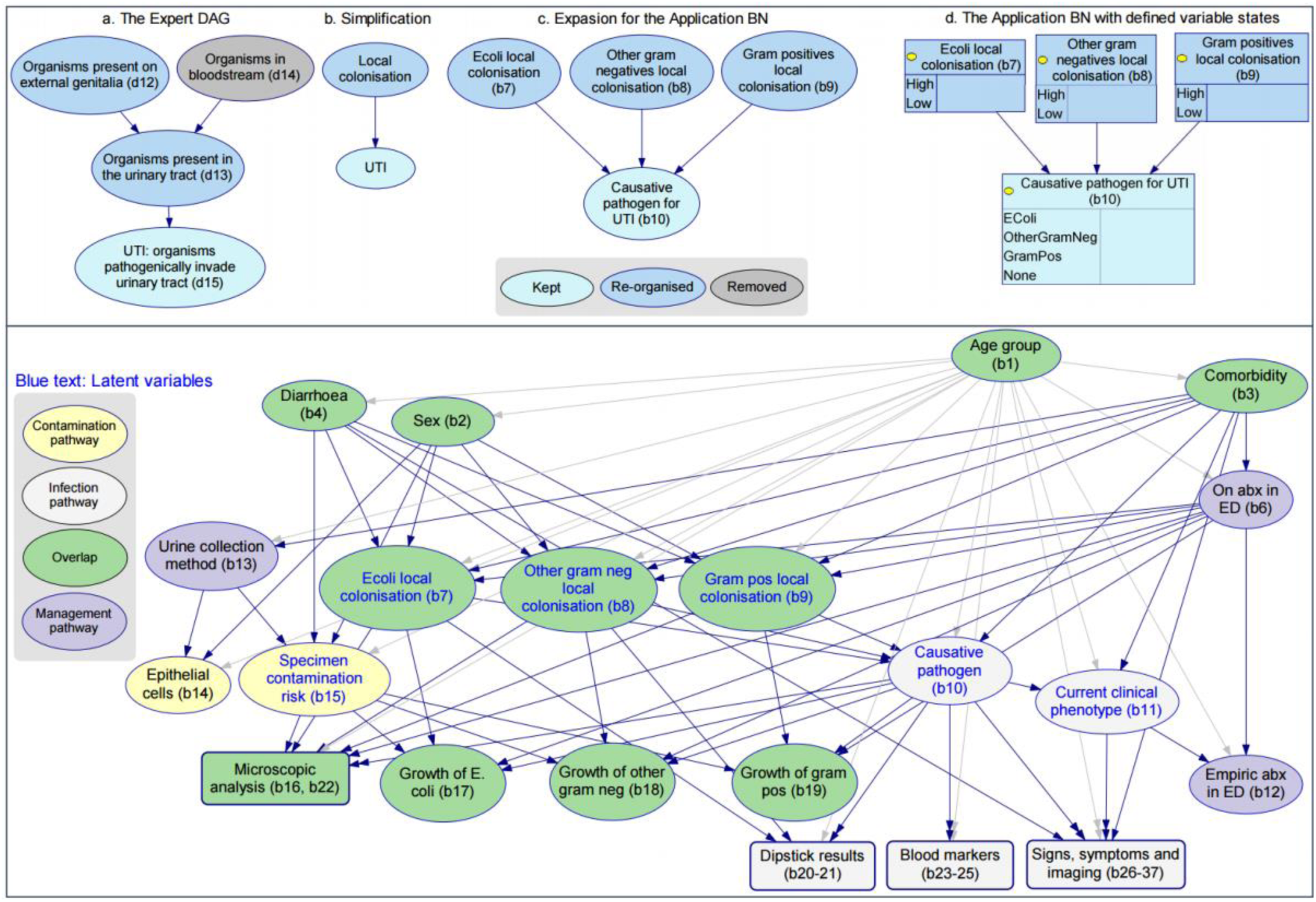
Top: An example of converting from the Expert DAG v11.1 to the Applied BN v2.2. Bottom: The high-level Applied BN structure. Additional file 4 includes a full list of differences between the two models. Additional file 5 presents the detailed structure of the Applied BN, in particular the local structure of submodels microscopic analysis, dipstick results, blood markers, and signs and symptoms (round box in the bottom panel), as well as the BN variable dictionary. The source model file for the Applied BN can be accessed via https://osf.io/8taqy/.

The Applied BN (Figure 2, bottom panel) comprises 36 nodes including 6 latent nodes, which can all be mapped to variables in the Expert DAG (see Additional file 4). Expert survey responses were collated (Additional file 6) to inform the CPT priors for the BN, which were further updated by training based on the prospective cohort data (as described in Section 2.3). Of note, the node ‘current clinical phenotype’ was introduced into the Applied BN as a summary node of patient presentation phenotypes after feedback from the expert validation workshop. This node is latent but was treated uniquely to provide a definition of current clinical phenotype that is independent of other latent factors in the model. In particular, a separate clustering was performed (using the EM algorithm) on the signs and symptoms, resulting in a grouping into three types, simply called “Type 1”, “Type 2” and “Type 3”, “Type 1” being systemic signs and symptoms predominant but mild urinary tract localising symptoms, “Type 2” being urinary tract localising symptoms predominant, and “Type 3” being abdominal pain predominant with minimal other symptoms. The clustering model was then used to determine each patient’s most probable clinical phenotype, and this information was added to the prospective cohort data in the form of an additional column and subsequently treated like an observed variable.

It is important to reiterate, by UTI, we mean the *existence* of UTI, which reflects the state of the world where a child’s urinary tract is infected by a pathogenic organism, and is only imprecisely defined. As a result, operational definitions of UTI and its causative pathogen vary across studies, and the definition is often incomplete (missing cases that we want to classify as UTIs). Evidence for UTI is indirect and comes from factors like cultures results and expert judgements, which is the way we approached it with our BN, leaving UTI as a latent variable and defined by its relationship with these other factors. The primary BN output is the **causative pathogen for UTI** (b10). Results from a 10-fold cross-validation show that the model predicts 68.0% of the presenting episodes in our cohort were UTIs, with IQR 67.2-68.9%. Specifically, this includes 40.2% *E.coli* UTI (IQR 39.3-40.8%), 11.6% other Gram negative UTI (IQR 11.6-11.9%), and 16.3% Gram positive UTI (IQR 14.9-18.0%). Figure 3 presents the Applied BN predictions for *E.coli* culture for every presenting episode, and compares these against their final laboratory results. The graphs represent four scenarios (a-d), each providing more information to the model than its preceding scenario. Namely, (a) provides the model with information on *basic demographics (age and sex) and clinical history (history of urinary tract pathology)*, (b) provides *(a) plus reported signs and symptoms*, (c) provides *(b) plus urine collection method and dipstick results*, and (d) provides *(c) plus all other available results (including urine microscopy and other clinical investigations).* The evaluation results show that the evaluation metrics (log loss and AUROC) improve as more evidence is available for a given child, especially if that evidence is sensitive and/or specific for UTI.

**Figure 3.**
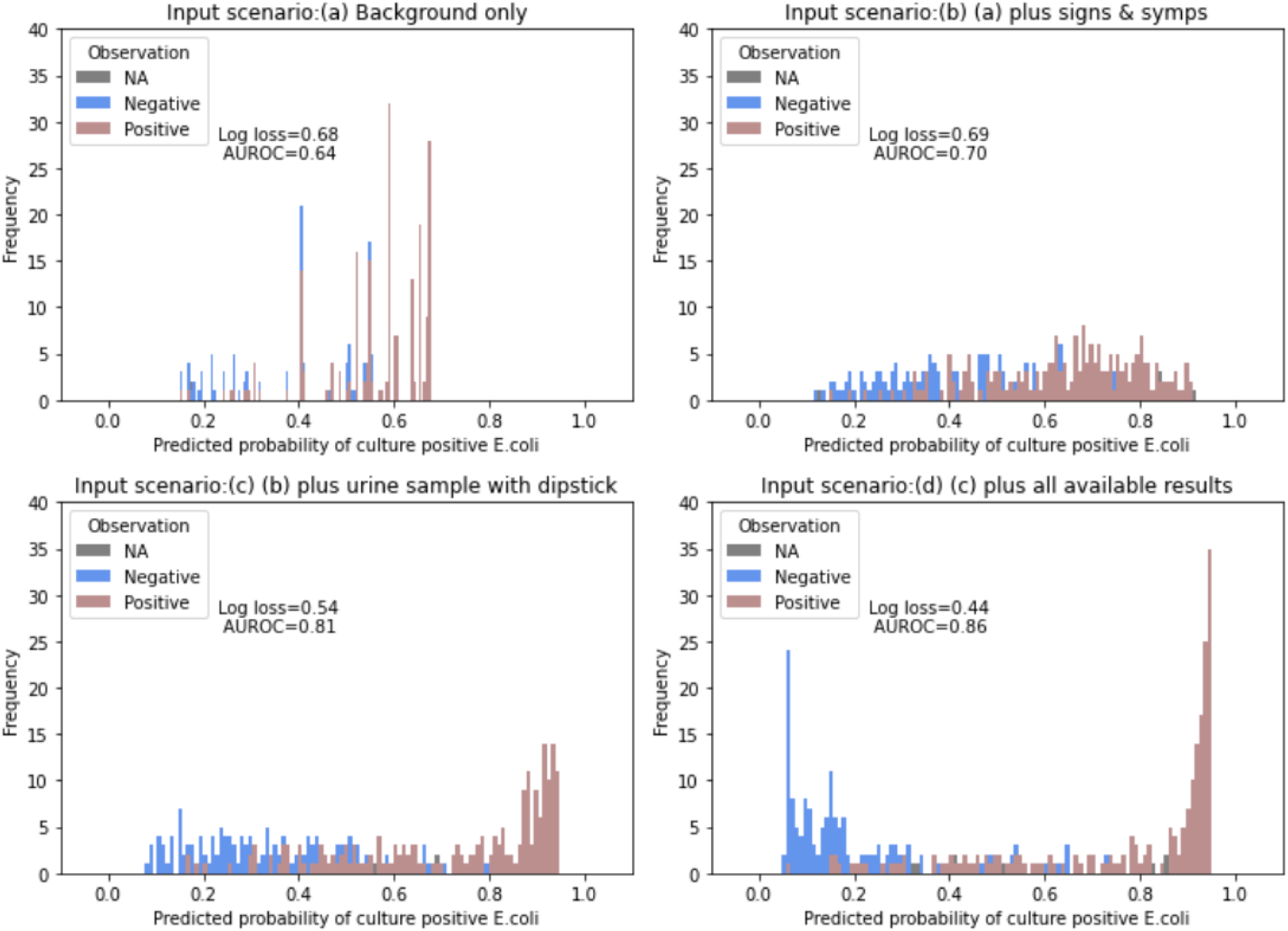
Applied BN v2.2 performance as compared with observations, with Log Loss and AUROC across four scenarios. Each panel presented the distribution of the Applied BN predicted probabilities of isolating *E.coli* from urine sample given available patient’s information under the specified scenario. The predicted probabilities were compared with the reported culture result of each patient, where brown, blue and grey indicated *E.coli* was isolated, not isolated and no data, respectively. Scenario (a): age, sex, history of UTI, urinary tract comorbidities. Scenario (b): scenario (a) + reported diarrhoea, urine tract pain or discomfort, abdominal pain, haematuria, foul smelling urine, respiratory symptoms, parent reported fever, temperature, irritability, lethargy, nausea/vomiting, poor oral intake. Scenario (c): scenario (b) + urine collection methods, urine dipstick results (leucocyte esterase & nitrite). Scenario (d): scenario (c) + urine microscopy (leucocytes, bacteria, epithelial cells), leucocyte and neutrophil count (from full blood count), C-reactive protein level and ultrasound result.

Two sets of parameters turned out to be very important in driving the primary target of the Applied BN (i.e., Causative pathogen, b10), namely, the probability of UTI in the prospective cohort (i.e., one minus the probability that Causative pathogen is none) and the pathogenicity (i.e., likelihood of causing disease and worsening illness) for each organism. Understanding the proportion of UTI and the pathogenicity of different organism groups is key as they determine how often a child would acquire UTI given local colonisation of an organism that is potentially pathogenic, which organism is more likely to be the causative pathogen when two or more organism groups co-colonise, and how likely the child would manifest as a more severe clinical case. These parameters were challenging to estimate as they were completely latent, hence we relied on expert opinion collected via a parameter survey as described earlier (Section 2.3). For the first of these parameters, the survey responses gave a mean estimate of 68% UTI among the study cohort (IQR 59-81%). Table 2 presents the survey outcomes for the second set of parameters on pathogenicity for each organism. We defined the pathogenicity of an organism as the propensity of the organism to cause UTI when an otherwise healthy child is colonised by that organism on the perineum or external genitalia. The survey elicited the pathogenicity of other Gram negative and Gram positive organisms as a numerical ratio relative to *E.coli*, on average, the responses suggest that *E.coli* and gram positive bacteria are very similar regarding their pathogenicity (1 and 0.98 respectively), and the other Gram negative bacteria is the most pathogenic (scored 1.35). Unlike other responses in the survey, the elicited responses for pathogenicity varied widely among experts. In Additional file 6, we provide a summary of responses to all survey questions.

**Table 2.** Expert survey outcome results of organism pathogenicity.

| Pathogenicity | Average | SD | Expert 1 | Expert 2 | Expert 3 | Expert 4 | Expert 5 | Expert 6 | Expert 7 | Expert 8 |
| --- | --- | --- | --- | --- | --- | --- | --- | --- | --- | --- |
| *E.coli* | 1 | 0 | 1 | 1 | 1 | 1 | 1 | 1 | 1 | 1 |
| Other Gram negatives | 1.35 | 0.53 | 1.5 | 2 | 0.8 | 1 | 0.75 | 1.75 | 1 | 2 |
| Gram positives (e.g., *Enterococcus*) | 0.98 | 0.91 | 0.5 | 0.2 | 0.5 | 1 | 0.375 | 2.75 | 0.5 | 2 |

Given the high level of variation in these survey outcomes, we therefore conducted sensitivity analyses by varying the prior CPT parameters for b10 by ±20%. In response, as shown in Figure 4, the predicted probability of UTI in our cohort of suspected UTIs ranged from 44 to 87%. *E.coli* is always predicted to be the most likely causative pathogen among the UTIs (39-64%), the relative attribution of other Gram negatives and Gram positives as the causative pathogen among the UTIs is sensitive to their pathogenicity, ranging from 12-29% and 22-32%, respectively.

**Figure 4.**
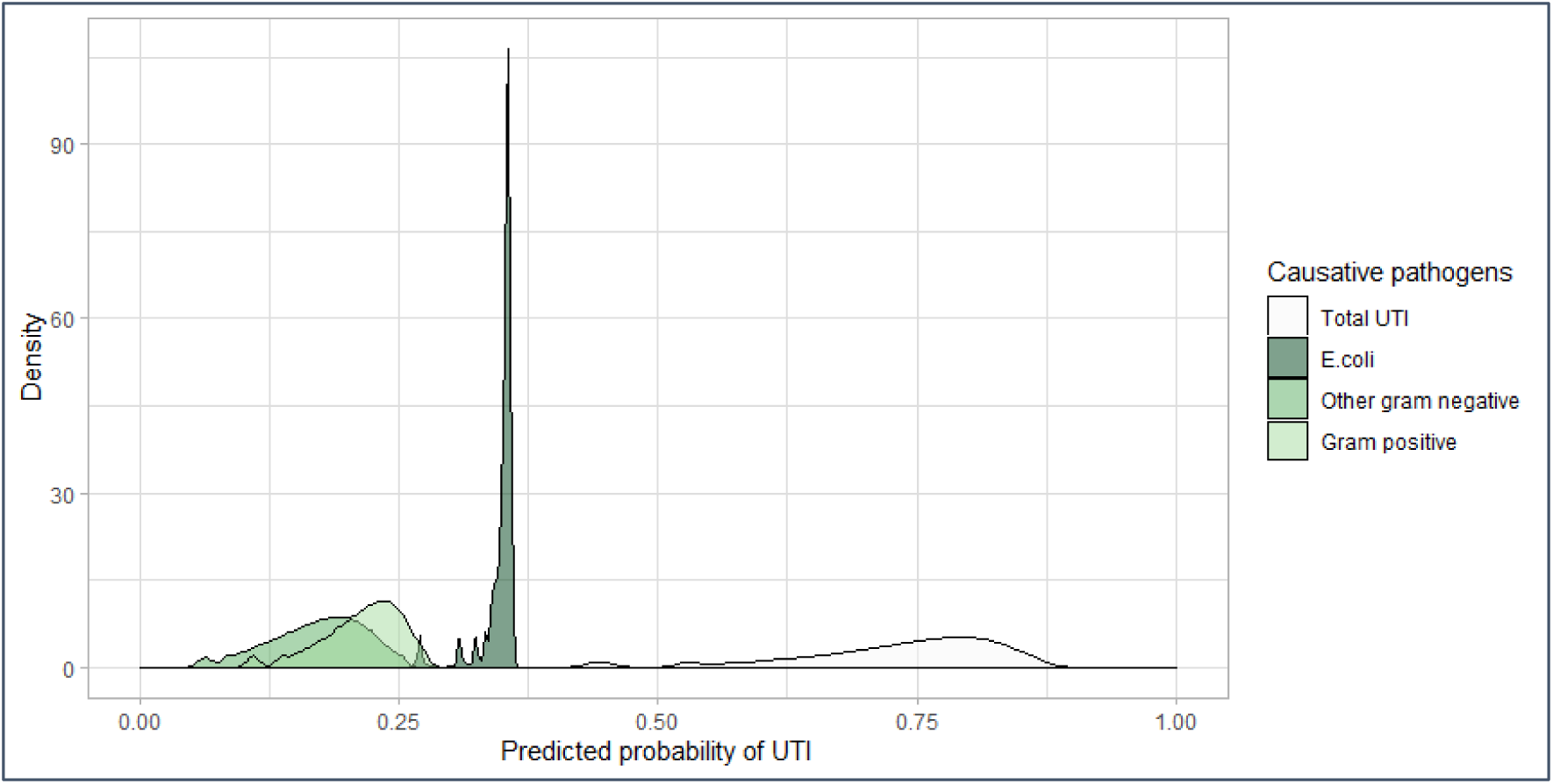
Sensitivity analysis on causative pathogens as the pathogenicity of different organism changes by ± 20%.

Figures 5-7 present three hypothetical clinical scenarios to illustrate how the Applied BN may be used for point of care decision support in the management of children with a suspected UTI. Predictions for each of the scenarios is shown branching conditional on various potential information and test results as they may become available over time. The scenario in Figure 5 presents an infant who is unable to communicate any localising symptoms. As information from the dipstick test, blood test and culture result become available, the BN’s predictions for the presence of UTI (and, if present, the associated causative pathogen) are updated accordingly. For example, when evidence from a dipstick result and CRP analysis are strongly indicative of UTI (i.e., “Nitrites detected” and “CRP 80”), a negative culture won’t exclude a UTI. Figure 6 presents a scenario in which UTI is always highly probable. Here, the presence or absence of comorbidities, temperature, dipstick nitrites and blood neutrophil levels only influence which causative pathogen is most likely. Finally, Figure 7 describes a child with no obvious localising symptoms, where a combination of test results can both rule in or rule out a UTI, as well as affect conclusions about the most likely causative pathogen.

**Figure 5.**
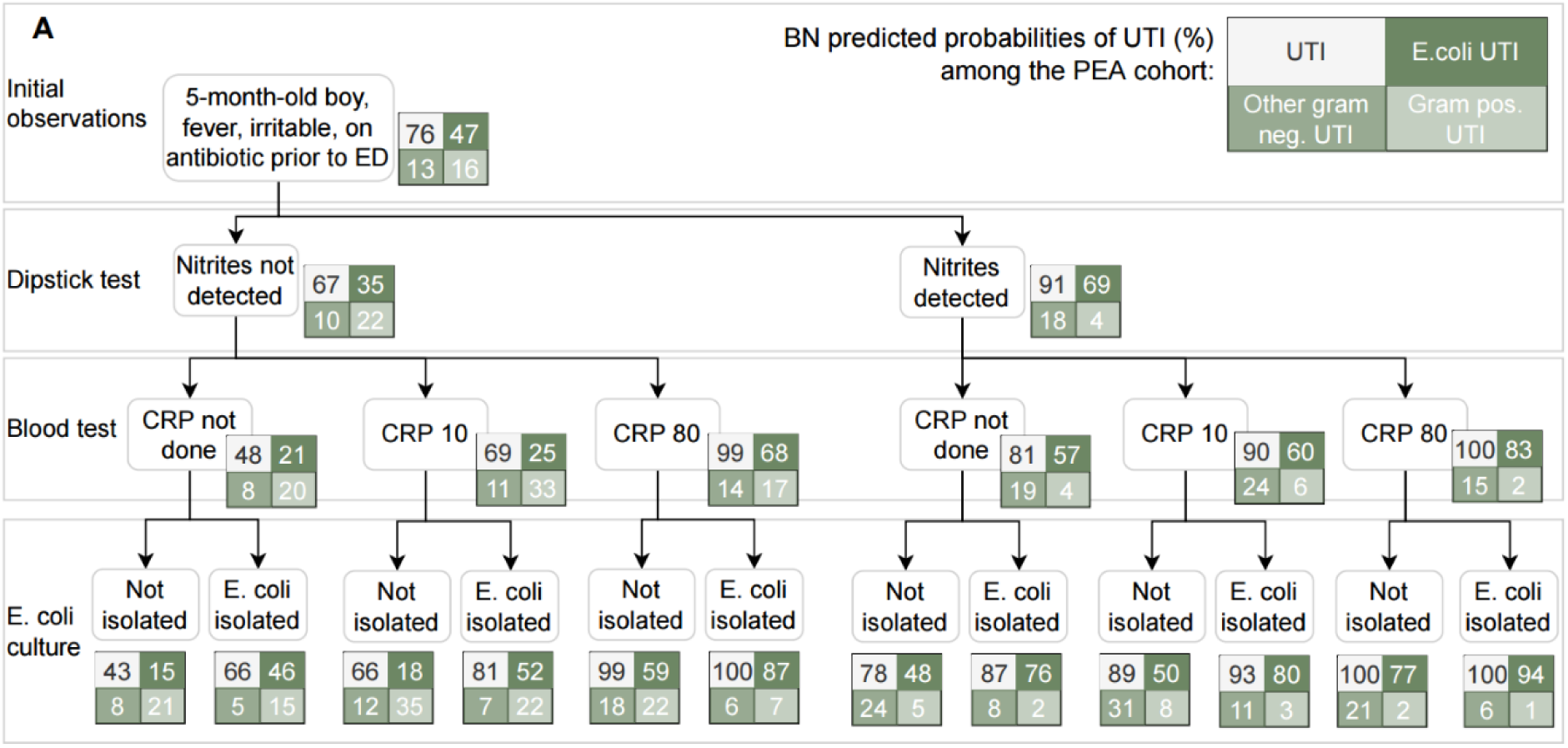
Predictions from the Applied BN under the clinical scenario A.

**Figure 6.**
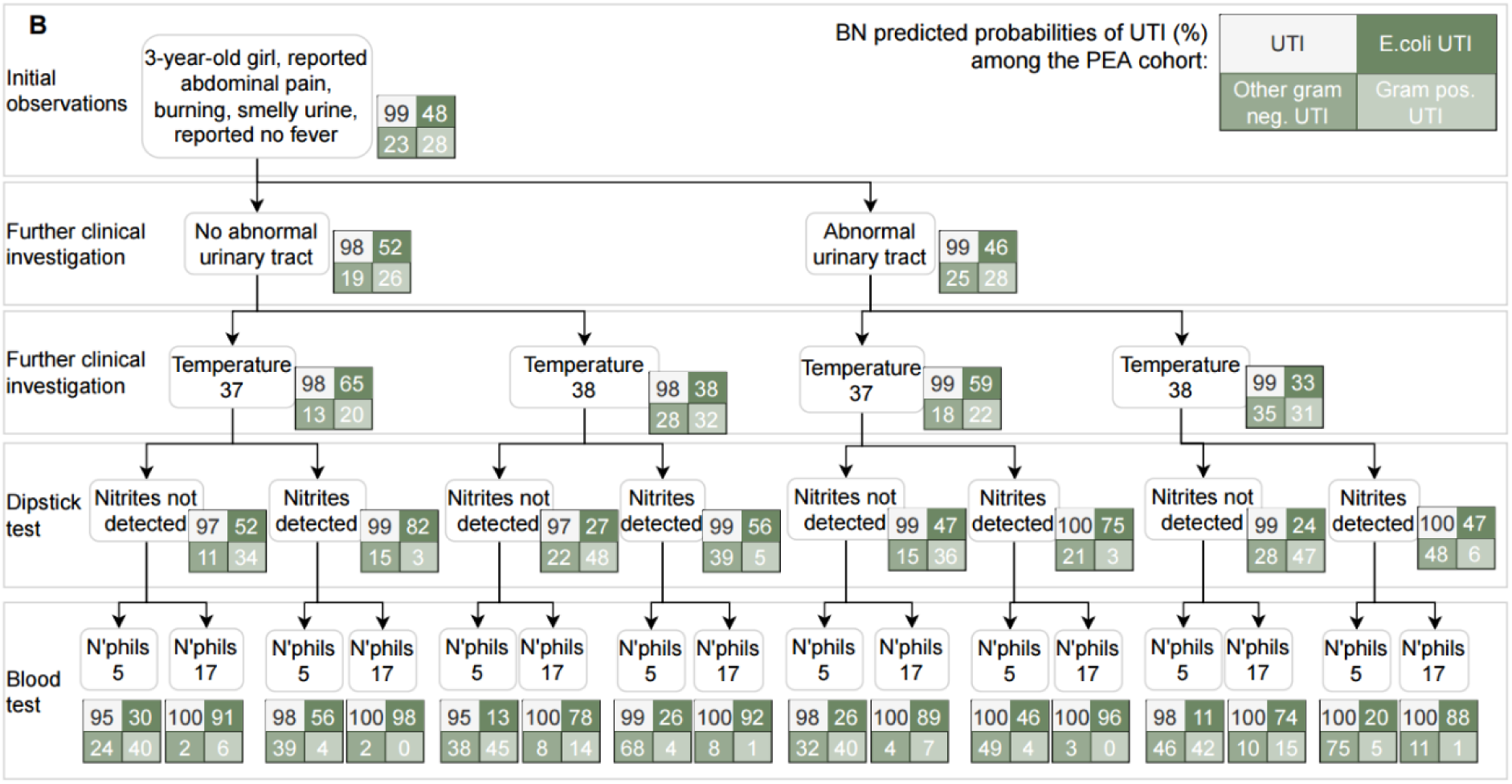
Predictions from the Applied BN under the clinical scenario B.

**Figure 7.**
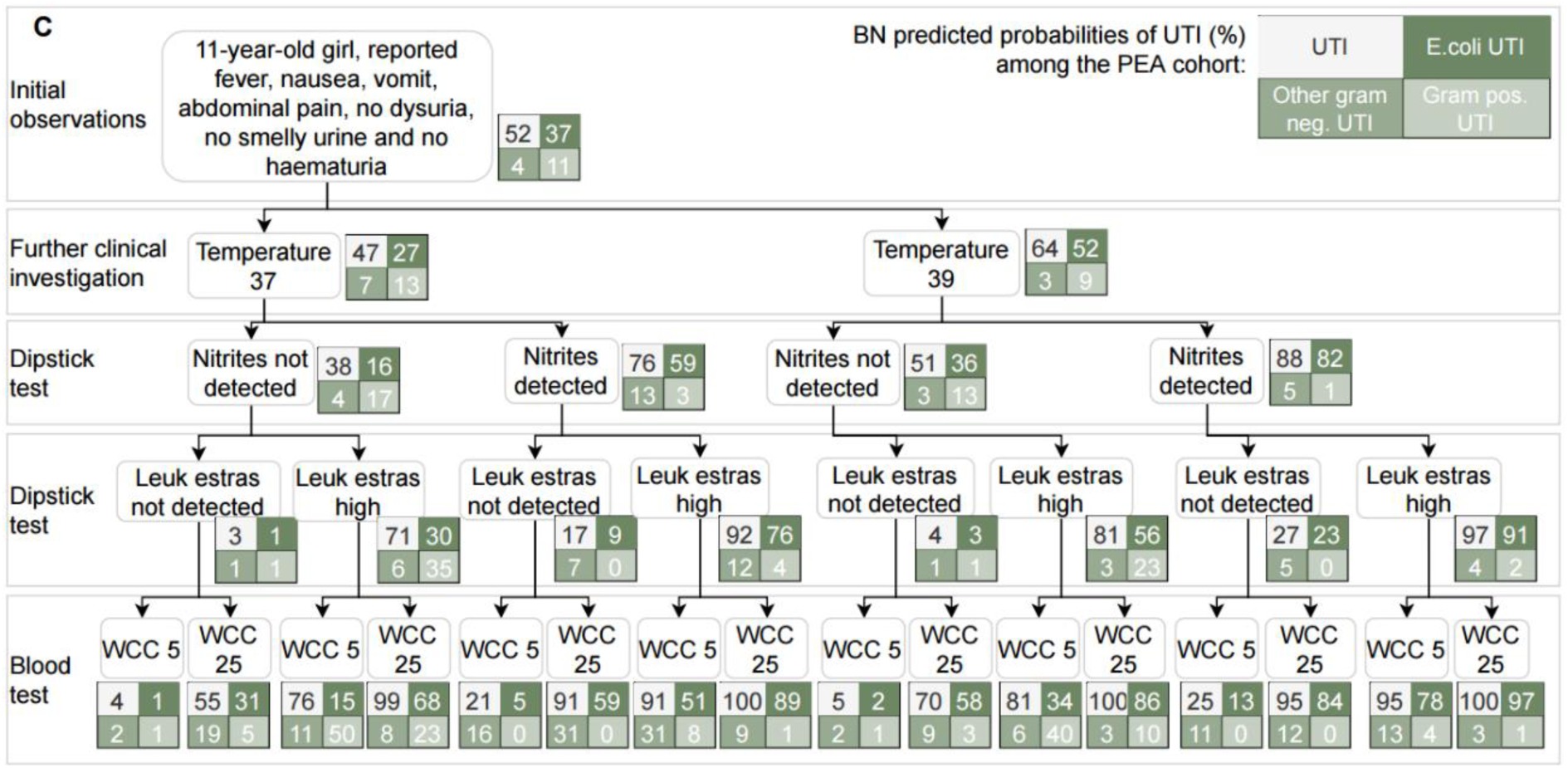
Predictions from the Applied BN under the clinical scenario C.

## 4. Discussion

Diagnosis and management of UTIs in children can be challenging due to the variability of clinical presentations and difficulties in obtaining a urine sample free from contamination. By mapping the causal pathways involved in this process through the development of an expert knowledge-derived DAG (the ***Expert DAG***, Figure 1 & Additional file 3), we have highlighted how the convergence of the causal pathways through sample collection and clinical diagnosis are key in creating this diagnostic challenge. Further to this, we have described which information may be available at different stages of the diagnostic and management process, and which additional evidence may be required to better understand the causal process. With the data collected from 431 episodes of suspected UTI in children, we converted the Expert DAG into a causal Applied Bayesian network model (the ***Applied BN***, Figure 2 & Additional file 5) to assess the probability of UTI (and if so the causative pathogen) among children with a suspected UTI.

The Applied BN achieved an excellent and stable performance in predicting *E. coli* culture results, with a mean AUROC of 0.86 and a mean log loss of 0.48 based on 10-fold cross-validation. We illustrated how the Applied BN could be implemented in practice as a clinical decision support tool using three hypothetical clinical scenarios.

### 4.1 The need for a better understanding of epidemiology and diagnosis of UTI

UTI epidemiology is primarily described based on urine culture results which are influenced by three causal pathways; (i) specimen contamination, where bacteria are introduced and not causative of the infection, (ii) clinical management, where empiric antibiotic exposure may suppress the bacteria causing an infection and/or specimen contamination, and (iii) the pathogenic causative organism of interest. Among the 431 episodes of suspected UTIs enrolled through the prospective cohort, after excluding 55 missing culture results, 60% specified growth of a bacterial organism, of which 90%, 7% and 6% were *E. coli*, other Gram negative bacteria and Gram positive bacteria, respectively. After the cohort data was used to train the Applied BN and thus interpreted under a causal framework, 69% of the study cohort were predicted to be UTIs, of which 57%, 16% and 27% were predicted to have been caused by *E. coli*, other Gram negatives, and Gram positive bacteria, respectively. The difference in the predicted distribution of causative pathogens by the causal model and crude microbiology data, which does not account for contamination and the effect of prior treatment, could have implications for antibiotic guidelines and urine culture reporting protocols.

More explicitly, the observed proportion of *E.coli* culture (54% of the overall prospective cohort) does not include all and only cases of UTI. The Applied BN suggests that: (i) specimen contamination results in 26% of urine culture isolates of *E.coli* being predicted to be non-UTIs and non-*E.coli* UTIs (i.e., false positives); and (ii) 84% of predicted *E.coli* UTIs reported growth of *E.coli*, implying a 16% false negative rate with a predicted 73% of prior antibiotic use. This concept is further described in the illustrative scenario of Figure 5, where for an irritable infant boy with fever and antibiotic use prior to ED, having no nitrites detected on urinary dipstick and with no CRP test performed, and where *E. coli* was isolated from the urine sample, the Applied BN predicts a 46% probability of this representing a *E. coli* UTI. In contrast, for the same child with nitrites detected on their urinary dipstick and with a CRP of 80 mg/L, the Applied BN predicts a 77% chance of a *E. coli* UTI, even if *E. coli* is not isolated from the urine sample.

When making decisions, clinicians are required to balance the risks associated with treatment based on a positive urine culture result that may not represent a UTI, against the risk of complications if UTIs are not adequately treated, particularly in neonates and young infants. By organising observable information within a causal DAG, we can highlight potential mediators, confounders and sources of selection bias and measurement errors (28). The prospective cohort study has mapped out the variation in the clinical picture of children investigated for a suspected UTI. Reported symptoms and urine analysis results differed greatly with age (Table 1), which likely represents an amalgam of children with and without UTI, and further highlights the need for decision support tools to distinguish between these groups. Mapping observable variables of the prospective cohort study cohort to the variables described in the Expert DAG, coupled with simplification and expansion, has enabled a quantitative model to be developed into a decision support tool (the Applied BN). Interpreting the observations available to clinicians under this causal framework may offer a clearer understanding of the clinical picture and provide robust assessments of the likelihood of UTI.

Organism-specific pathogenicity needs to be better understood to improve the diagnosis of the causative pathogen for each UTI. Based on our experts’ survey responses (Table 2), we assumed that non-*E. coli* Gram negative bacteria have the greatest pathogenicity in the current model, while *E.coli* and Gram positive pathogens have similar and lower pathogenicity. These assumptions can have implications for the model predictions. An example can be found in the scenario of Figure 6, where, for a 3-year-old girl with reported abdominal pain, smelly urine, burning, no parent reported fever but a recorded temperature of 38 degrees, nitrites not detected on urinary dipstick and blood neutrophil count of 5 x10^9^/L, the Applied BN predicts other Gram negative bacteria as the most likely causative pathogen, regardless of whether the child has any history of urinary tract pathology. However, the survey outcomes indicated a high level of variation regarding the relative expert-derived pathogenicity of different organism groups. This was especially relevant for Gram positive organisms where growth is often attributed to contamination and the ability of some organisms to cause UTI may be disputed. As a result, the Applied BN only demonstrates there is a potential to differentiate causative pathogens for UTI like non-*E.coli* Gram negative and Gram positive bacteria, based on assumed pathogenicity and current data. While it’s not yet ready to be used for differentiating pathogens, it does suggest a way forward in understanding the organism-specific pathogenicity.

### 4.2 Learnings from the modelling process

The Expert DAG demonstrated that specimen contamination risk, propensity to develop complications and an organism invading the urinary tract system were the key latent concepts that concerned clinical teams. Importantly, superficial colonisation of the perineum/ genitalia lies on the causal pathways mediating both invasion of the urinary tract system and specimen contamination which converge at urine culture, the only point at which either of the two pathways is typically observed. We therefore chose to model these variables explicitly in the Applied BN as pathogen-specific ‘local colonisation’ (b7-b9), ‘causative pathogen’ for UTI (b10) and ‘specimen contamination risk’ (b13), despite the challenges of parameterising these latent nodes. We addressed this challenge by designing survey questions to elicit estimates of relevant parameters from the domain experts. In some cases, even those expert elicited responses were inconclusive (namely, the organism-specific pathogenicity) and in those cases we conducted sensitivity analyses to ensure the implications and limitations of the uncertain parameters were recognised (as discussed for pathogenicity in the previous section).

Where possible, the Expert DAG was causal and comprehensive of the problem domain rather than constrained by variable observability or data availability. This allowed it to be used as an accurate representation of expert knowledge, enabling the use, adaptation and extension by the core research team and external researcher. By documenting the detailed steps of the conversion from the Expert DAG to the Applied BN (Additional file 4), we established a methodological framework that can be generalised beyond the UTI problem domain. Decisions to keep, remove or add variables in the applied model should be driven by a well-defined modelling purpose, matched to the availability and quality of data, and technical efficiency (such as reducing the number of latent nodes, or reducing the complexity of the variable relationships).

Like most complex modelling work, our variable selection, structure development, parameterisation and evaluation processes were iterative. The communication between modellers and the domain experts played an important role in this project, which required both parties to make efforts to understand each other’s expertise and language. Medical education focuses on the pathophysiology of disease, where factor ‘X’ predisposes to outcome ‘Y’. However, in practice, clinicians are more experienced in using rule-based flow charts and decision trees to aid in management, which depict ‘if [specific signs and symptoms], then perform [this test]; if [this result] then commence [this treatment]’. The creation of an expert-derived DAG required clinicians not only to revisit the concepts of the causal effects of each variable and their direct influence on another, but also to depart from the concept of a graph reflecting a sequence of steps or yes/no questions to observations, and instead that one may have the real outcome of interest (e.g. the *existence* of UTI) existing as a latent node in the body of the model, influencing the observable nodes that appear below it. Similarly, the concept of a latent node was challenging, given that clinicians typically work on the premise that they have the correct (i.e., ‘true’) diagnosis that informs their treatment decisions. While clinicians are certainly familiar with the related concepts of false positives and negatives, the extension to latent nodes was not straightforward and required more guidance from the core research team. The creation of a DAG highlights the fact that some important variables will always remain unobserved and therefore uncertain; although evidence may be accumulated to increase certainty about the presence or absence of infection, in reality infection can only ever be inferred and never directly observed. Becoming comfortable with these concepts enabled the experts to create the elicited DAG and understand its utility in clinical practice in the form of a BN.

### 4.3 Study limitations and future research

The prospective study cohort aimed to describe participants <13 years of age who presented into the ED and were prescribed antibiotics for a suspected UTI. With these criteria the data used to develop the models and resultant models likely represents patients that have more severe and complex disease and a greater risk of hospitalisation and antimicrobial resistance than those presenting for a UTI within the community. In other words, selection bias may be generated at the time patients were screened for eligibility and recruited for data collection, limiting the use of the Applied BN to the same cohort. Microbiology data obtained as part of the prospective study cohort was limited. The distribution of pathogens was likely representative, however, there were a small number of samples that isolated non-*E. coli* Gram negative bacteria and Gram positive bacteria. This required a broad pathogen grouping which may have included bacteria with greatly differing uropathogenic characteristics, as a result, only a limited understanding of how clinical and laboratory variables can help differentiate causative pathogens was developed. Further to this, a greater understanding of colonisation, infection and bacteria specific pathogenicity in the urinary tract is required to further the development of this model, yet much of this information is debated widely in the scientific community (29). The Applied BN briefly describes the empiric antibiotic prescribing patterns within the ED where 62% and 38% of described antibiotics prescriptions were narrow and broad spectrum, respectively. It is intended this model will be expanded with additional information on antimicrobial susceptibility profiles to evaluate the appropriateness of empiric antibiotic prescriptions for a range of causative pathogens.

With a richer dataset, our models could benefit from further development that could provide predictions for a broader scope, for example, incorporating how decisions were made on collecting urines and conducting blood tests, as well as potential other diagnoses other than UTI. We provide this model in its current updatable form for further parameterisation, validation, and extension by external and future researchers. The model can be adapted across a range of laboratories, hospitals and patient populations, and we anticipate this framework will aid the interpretation of culture results, the diagnosis of UTIs, and choice of antibiotic prescription, and can be incorporated into routine clinical pathways with the overall goal of improving patient outcomes and reducing inappropriate antibiotic use in children. To our knowledge this is the first causal BN for UTIs in children; we believe it serves as an exemplar for the creation and use of causal model-based decision support tools across a broad range of infectious disease problems.

## Data Availability

All source models produced in the present study are available online at https://osf.io/8taqy/.

## Supplementary material

Additional file 1: Schematic of participant enrolment and data collection

Additional file 2: Parameterisation survey questions

Additional file 3: The Expert DAG variable dictionary

Additional file 4: List of changes when converting the Expert DAG to the Applied BN

Additional file 5: Full structure of the Applied BN and the BN dictionary

Additional file 6: Parameterisation survey responses.

## Declarations

### Ethics approval and consent to participate

Ethics approval was granted by the Child and Adolescent Health Service Human Research Ethics Committee (EC00268). Informed consent was provided by the legal guardian of each participant.

### Consent for publication

All authors provided consent for the publication of this work.

### Availability of data and materials

All source models and associated dictionaries are accessible as additional files to the manuscript, and via our Open Science Framework page, https://osf.io/8taqy/.

### Authors’ contributions

TS and YW initiated the project. YW designed the project. JAR, AJC, DF and TR led the data collection. YW and JAR led data analysis and interpretation. YW, SM, JAR and TS led the DAG development. SM and YW lead the knowledge elicitation and the BN modelling, and model evaluation activities. TS, AJC, DF, AOM, PI, MLB, CB, NGL, TR and PW participated in the development of models as domain experts. JAR and YW lead the manuscript writing, and TS, SM and AOM substantially contributed to the writing. All authors have contributed to the writing and reviewed and approved the final manuscript submission.

## Acknowledgements

We acknowledge all enrolled participants and their family for contributing data to this research. We acknowledge research nurses Sharon O’Brien, Lisa Properjohn, Mel Dowd, Katy Whitten and Jacq Noonan at Perth Children’s Hospital, emergency department for data collection.

## Additional file 1: Schematic of participant enrolment and data collection

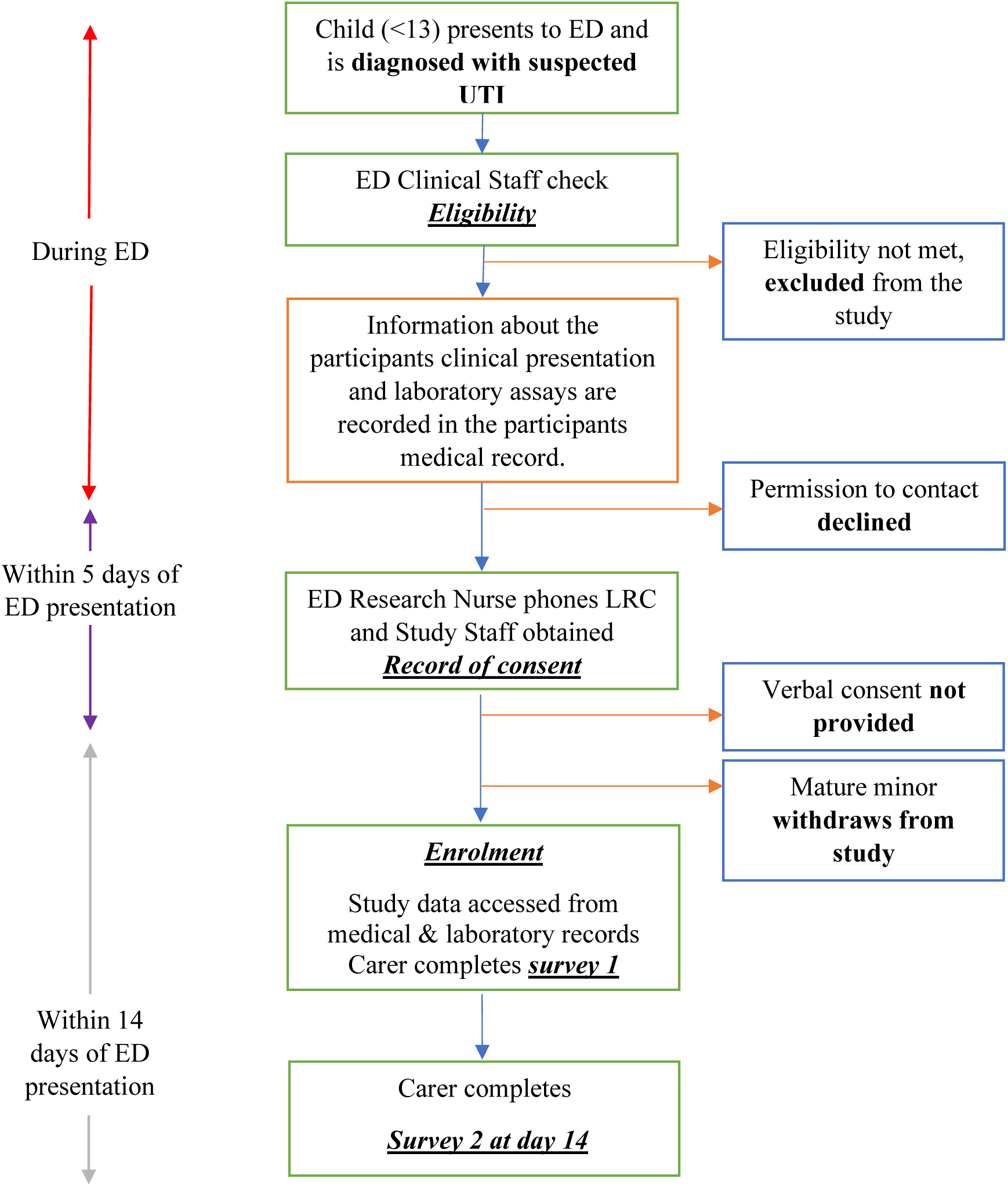

## Additional file 2: The parameterisation survey questions

In this document we provided all survey questions used for the elicitation of parameters.

### Q1. Risk of specimen contamination

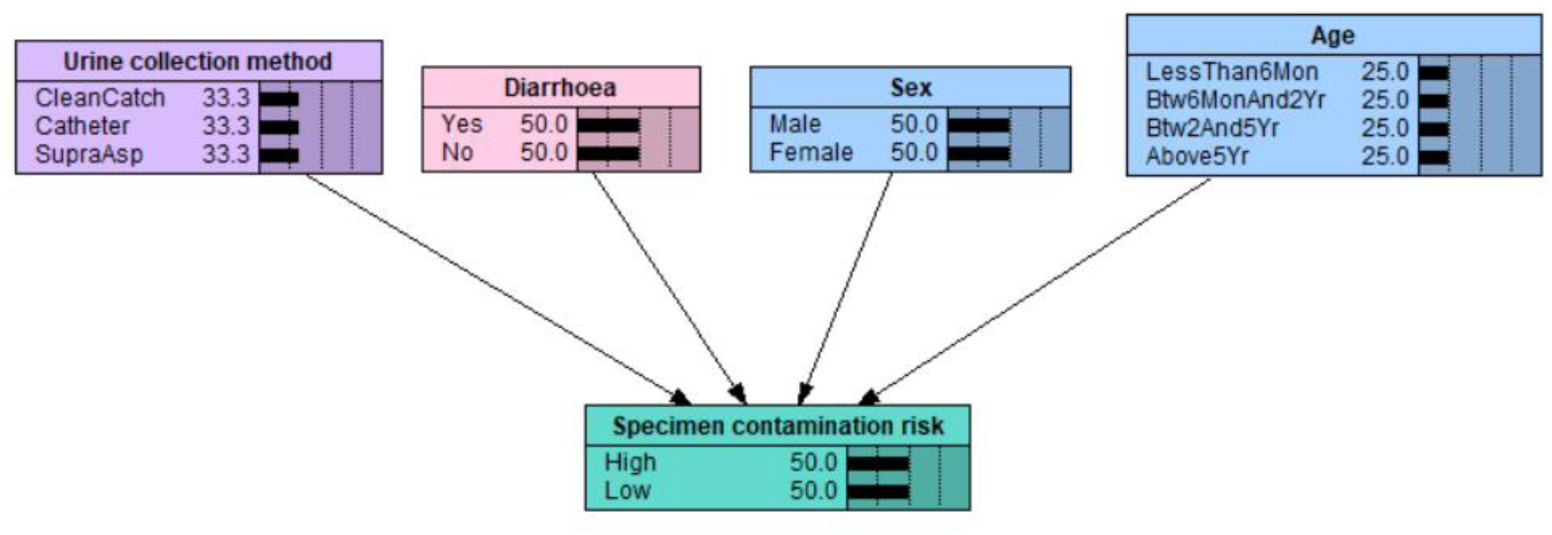

Consider the risk of a non-causative organism/s entering the urine specimen during the specimen collection process. In the model (as shown in the above figure), the **risk of specimen contamination** is influenced by **age**, **sex**, presence of **diarrhoea**, and **urine collection method**. Assuming *the same* colonisation status of each child’s perineum/ external genitalia (i.e., type and density of organisms), how do the following factors increase or decrease the risk of specimen contamination from the baseline (as specified below)? E.g., x0.3, x2, x10, etc.

1a. **Age** and **sex**, assuming **clean catch** as the method of specimen collection.

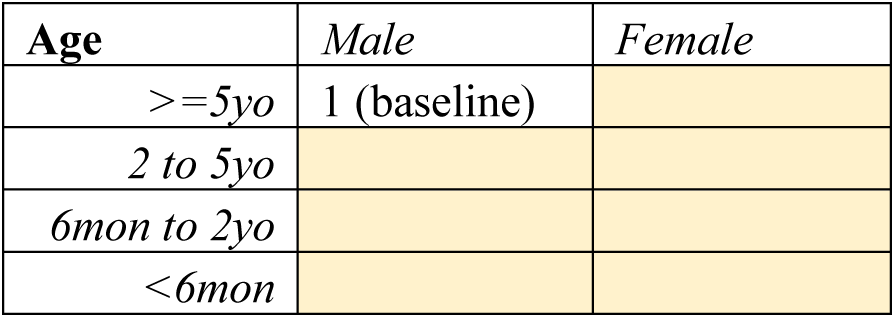

1b. Presence of **diarrhoea**, assuming **clean catch** as the method of specimen collection.

|  |  |
| --- | --- |
| Absence | 1 (baseline) |
| Presence |  |

1c. **Urine collection method**

|  |  |
| --- | --- |
| Supra aspirate | 1 (baseline) |
| Catheter |  |
| Clean catch |  |

1d. Any further comments?

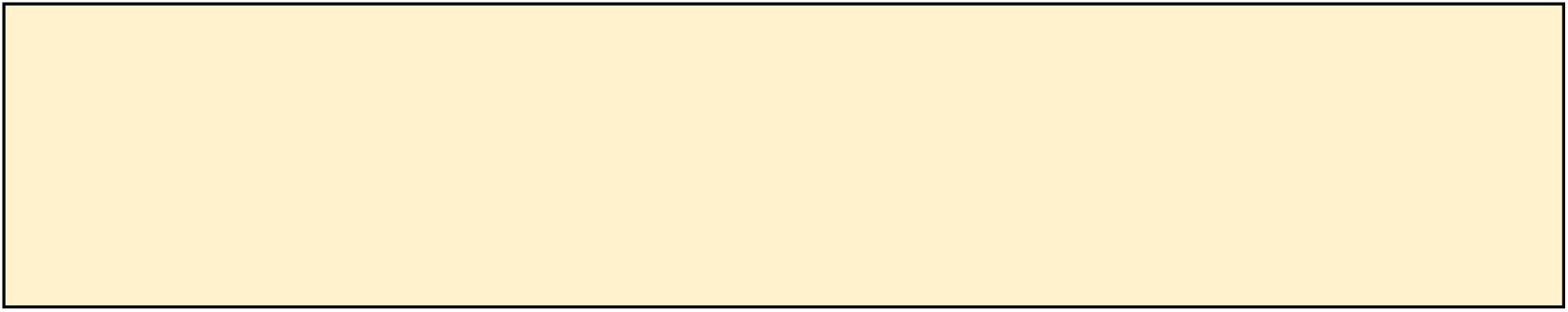

### Q2. Propensity to UTI progression

Consider a child’s risk of progressing to more severe disease manifestations given they have a UTI, e.g., developing kidney infection, or experiencing worsening severity of local or systemic inflammatory response, which can be further broken into two concepts: the **speed of progression**, and the **susceptibility to severity** – illustrated using the diagram below. Please note that these curves are illustrative, not exact.

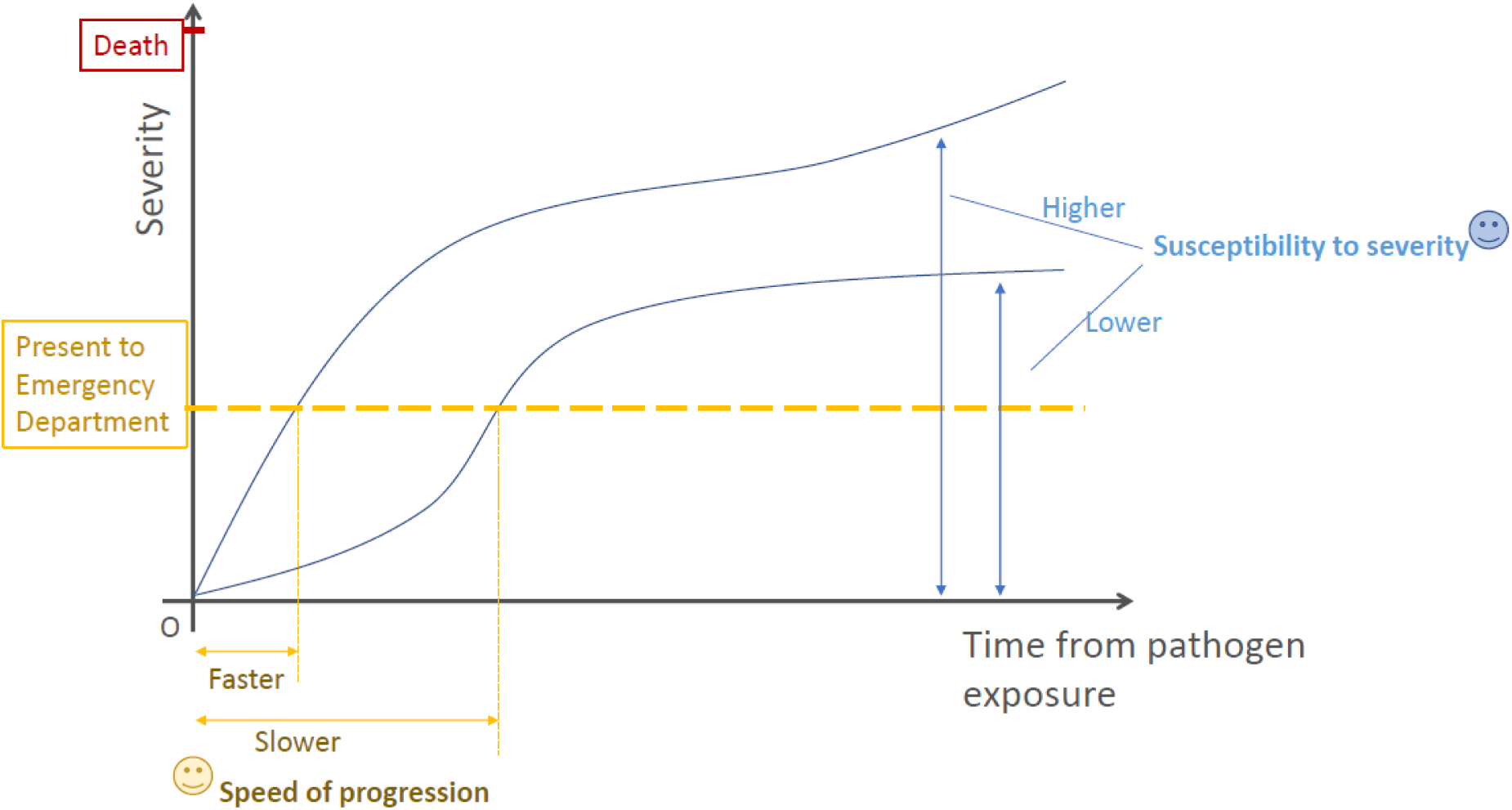

In the model, both the **speed of progression** and **susceptibility to severity** may be influenced by **age** and **UTI-relevant comorbidity** (such as VUR/anatomical abnormalities of the urinary tract). We now ask a series of questions on these two concept variables. Please provide your min, max, and best guess estimates for each question. Please note that the “min/max” should be plausible lower or upper values, e.g., 95th percentiles, not the extreme recordable value.

#### 2a. Speed of progression

Assuming a baseline speed of progression (as specified below), how do the following factors increase or decrease the baseline? E.g., x0.3, x2, x10, etc.

**Age**

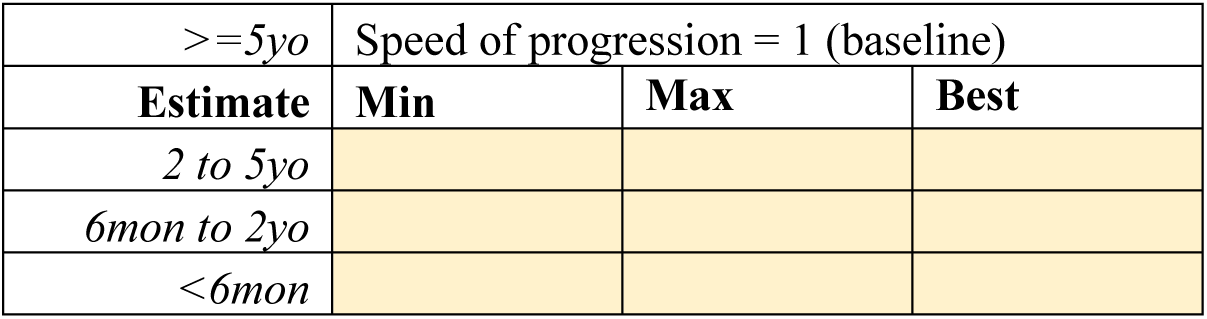

**UTI-relevant comorbidity**

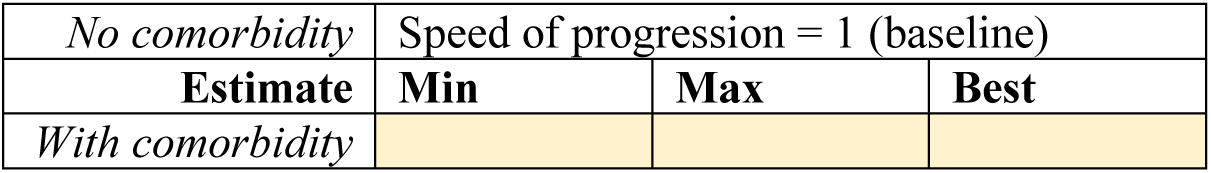

Any further comments on the speed of progression?

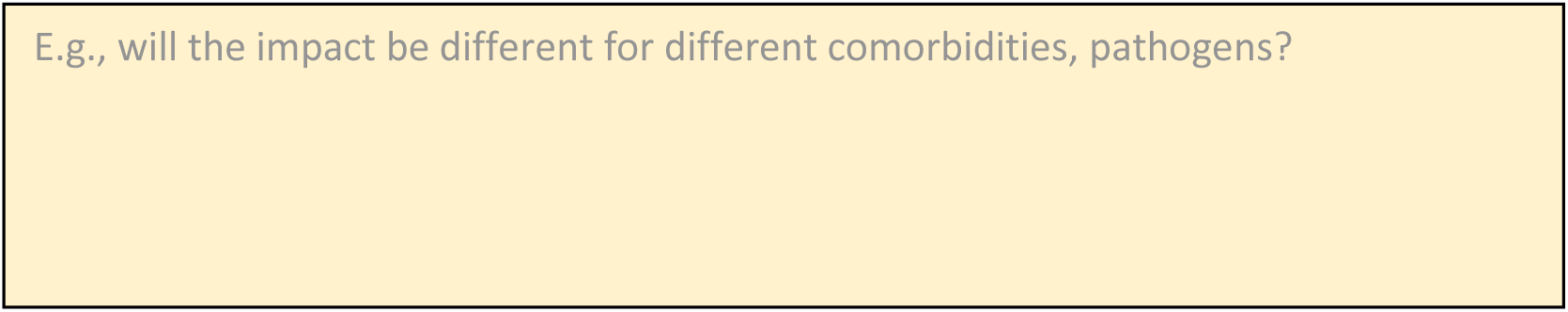

#### 2b. Susceptibility to severity

Assuming a baseline susceptibility to severity (as specified below), how do the following factors increase or decrease the baseline? E.g., x0.3, x2, x10, etc.

**Age**

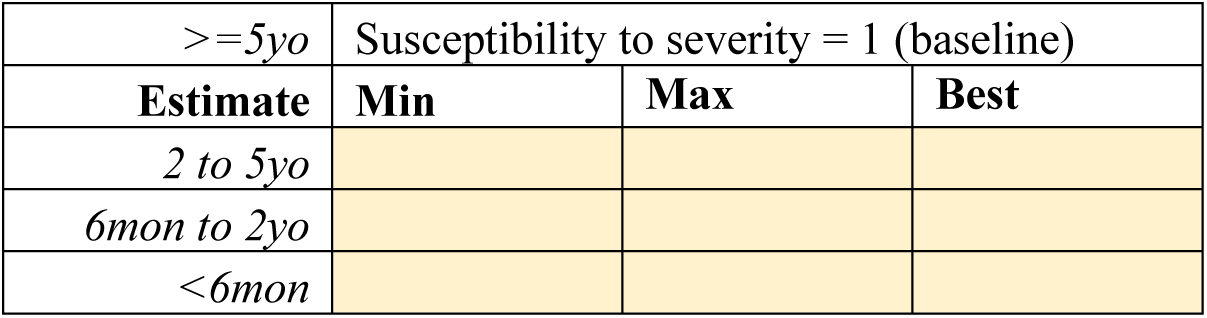

**UTI-relevant comorbidity**

|  |  |  |  |
| --- | --- | --- | --- |
| *No comorbidity* | Susceptibility to severity = 1 (baseline) |  |  |
| **Estimate** | **Min** | **Max** | **Best** |
| *With comorbidity* |  |  |  |

Any further comments on the susceptibility to severity?

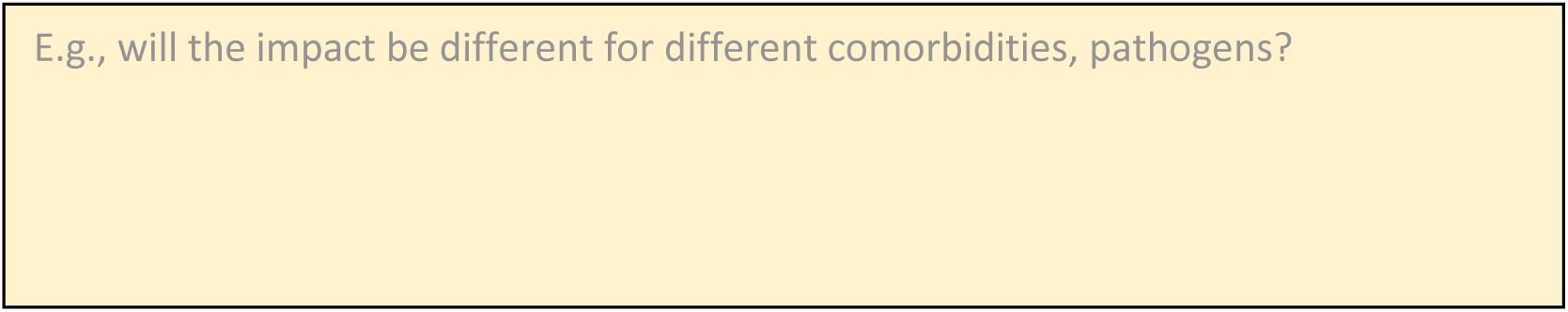

### Q3. Causative pathogen for UTI

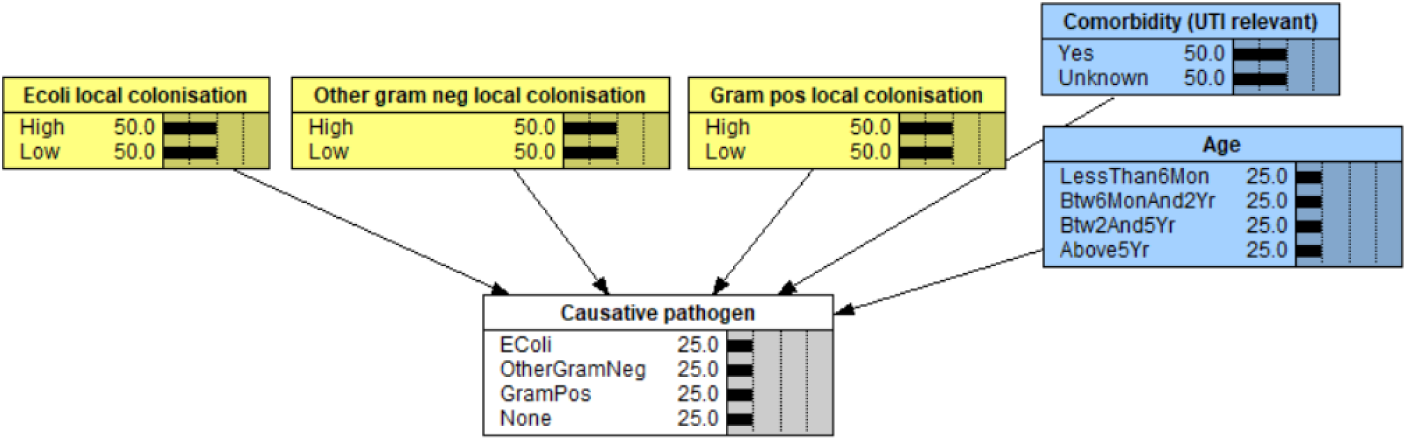

Colonisation of the perineum/ external genitalia by bacteria is assumed to predispose children to urinary tract infection (UTI). In the model, the probability of UTI with each causative pathogen (shown as **causative pathogen** in the above figure) is influenced by **age**, **local colonisation**, and **UTI-relevant comorbidity** (such as VUR/anatomical abnormalities of the urinary tract).

3a. Consider the PEA cohort, we enrolled children who presented to the Emergency Department (ED) at Perth Children’s Hospital and were managed for presumed UTI (with an antibiotic prescription in the ED and a urine sample sent for laboratory investigation). These patients typically underwent urine dipstick in the ED. What do you estimate <u>the probability</u> (min, max, best guess) of true UTI in this cohort (prior to seeing the laboratory culture result)? Please note that the “min/max” should be plausible lower or upper values, not the extreme recordable value.

|  | True UTI, in % |  |  |  |  |  |
| --- | --- | --- | --- | --- | --- | --- |
| Age | Boy |  |  | Girl |  |  |
| Estimate | Min | Max | Best | Min | Max | Best |
| $\geq 5yo$ | | | | | | |
| 2 to 5yo |  |  |  |  |  |  |
| 6mon to 2yo |  |  |  |  |  |  |
| <6mon |  |  |  |  |  |  |

3b. In the case of an otherwise <u>healthy child</u> with colonisation of the perineum/ external genitalia by all the following three groups of organisms: E.coli, other gram negatives, and gram positives. Note, we refer to gram positives that can potentially cause UTI, such as Enterococcus, rather than gram positives like Staph epidermidis which are unlikely cause UTI.

|  |  |  |
| --- | --- | --- |
|  | Consider the E coli pathogenicity as baseline, could you please indicate the relative pathogenicity of others? E.g., x0.5, x3, etc. | Please comment if different pathogens affect the **speed of progression** and **susceptibility to severity** differently? If so, how? |
| E.coli | 1 (baseline) |  |
| Other gram negatives |  |  |
| Gram positives (e.g., Enterococcus) |  |  |

3c. Any further comments?

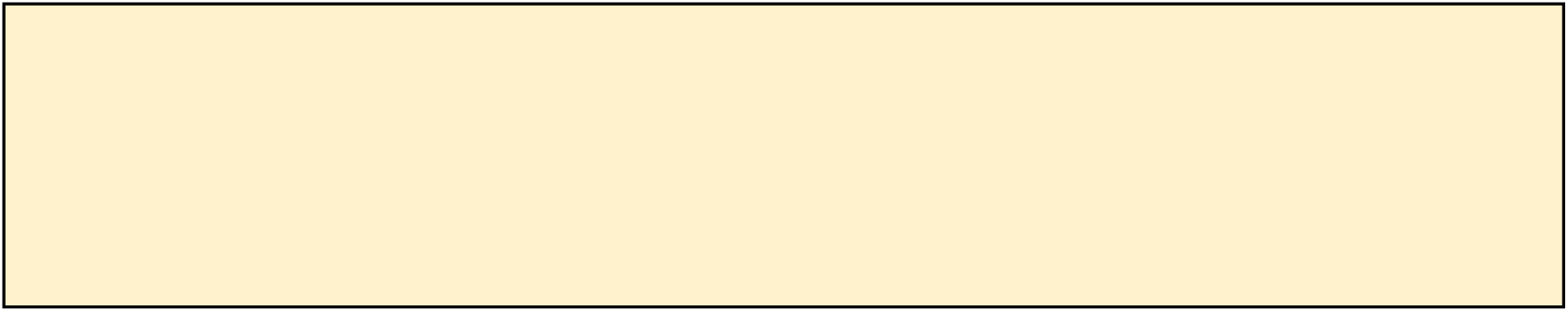

### Q4. Impact of exiting antibiotic use

4a. For modelling purpose, we have grouped antibiotics into two groups: narrow and broader, could you please review this grouping and suggest if any antibiotic should be grouped differently? Please feel free to add new group/s.

**Narrow:** Amoxicilin, Amoxicillin + clavulanic acid, Trimethoprim, Trimethoprim + Sulfamethoxazole, Benzylpenicillin, Cefalexin, Cefazolin, Co-trimoxazole, Erythromicin

**Broader:** Amikacin, Cefepime, Cefotaxime, Ceftazidime, Ceftriaxone, Ciprofloxacin, Colistin, Ertapenem, Gentamicin, Meropenem, Moxifloxacin, Nitrofurantoin, Norfloxacin, Piperacillin + Tazobactam, Tazocin, Tobramycin, Vancomycin

Any further comments?

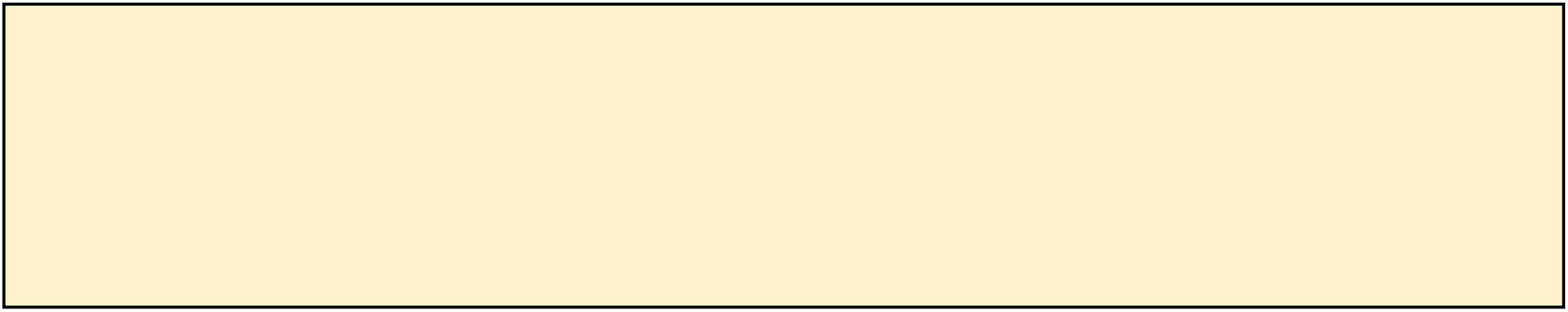

4b. Given a UTI, the successful detection of the causative pathogen of the UTI in laboratory can be influenced if the patient has been on antibiotic when they came to the ED where the urine sample was taken. Presumably this is largely affected by the antimicrobial susceptibility pattern of the pathogen which can variable by different subgroups, so please consider an average community-acquired case in 2019-2020.

Under the following scenarios, please provide your min, max, and best guess estimates for each question. Please note that the “min/max” should be plausible lower or upper values, not the extreme recordable value. (Please feel free to refer to your experience of treating UTI in adults.)

Consider a UTI caused by E.coli

|  | Probability of positive culture of E.coli |  |  |
| --- | --- | --- | --- |
| Estimate | Min | Max | Best guess |
| *Not on abx* |  |  |  |
| *On narrow abx* |  |  |  |
| *On broader abx* |  |  |  |
| *Pls feel free to add more groups* |  |  |  |

Consider a UTI caused by other gram negative bacteria

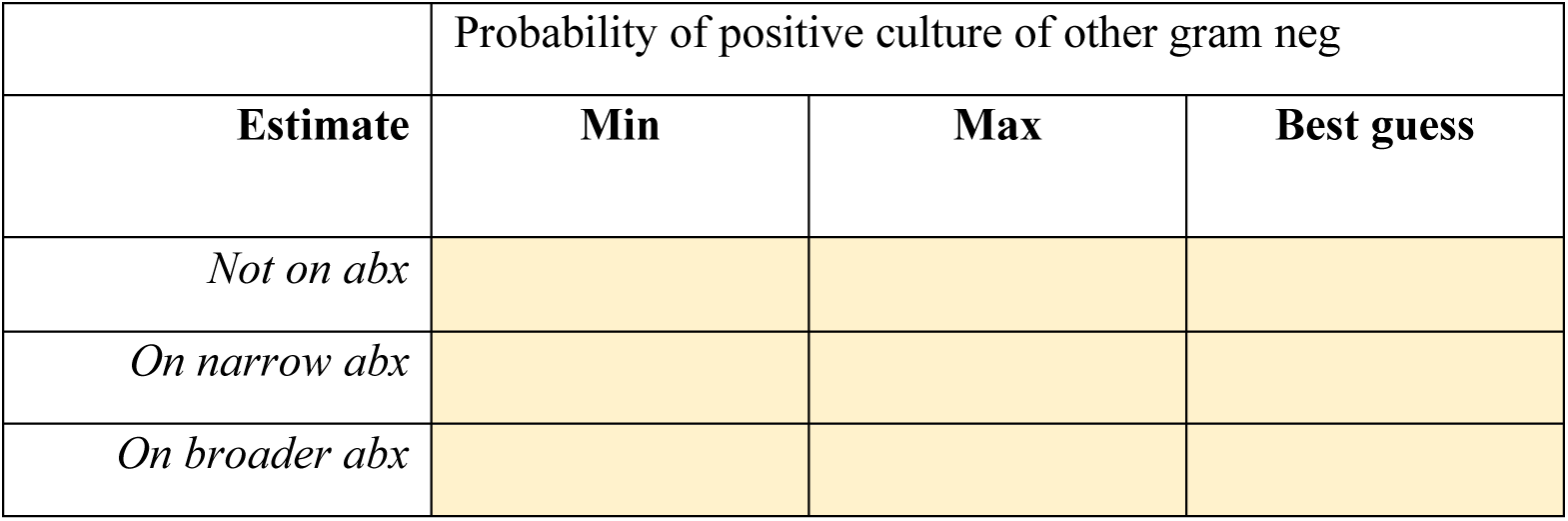

Consider a UTI caused by gram positive bacteria (e.g., Enterococcus)

|  | Probability of positive culture of gram pos |  |  |
| --- | --- | --- | --- |
| **Estimate** | **Min** | **Max** | **Best guess** |
| *Not on abx* |  |  |  |
| *On narrow abx* |  |  |  |
| *On broader abx* |  |  |  |

Any further comments?

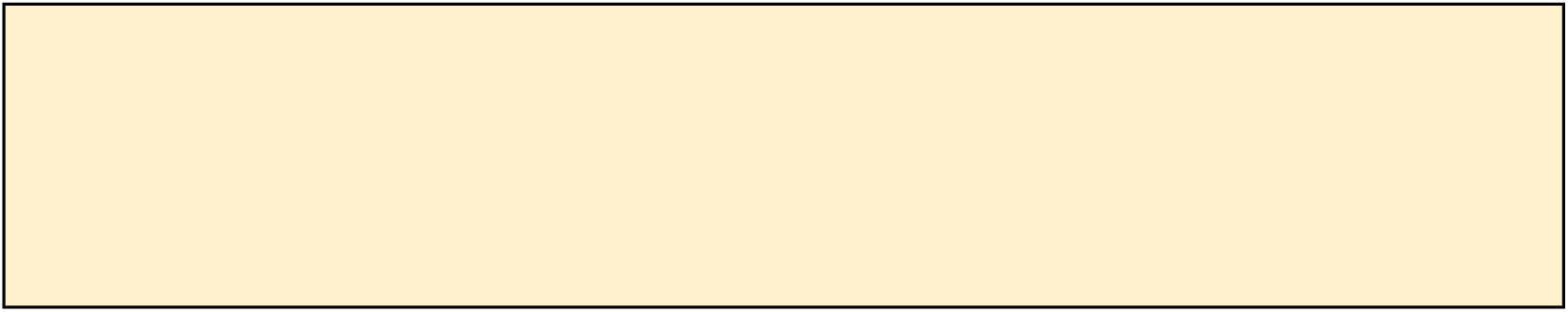

## Additional file 3: The Expert DAG dictionary

In this document we provided the structure of the Expert DAG v11.1 (Figures C1) and its dictionary (Table C1).

**Figure C1.**
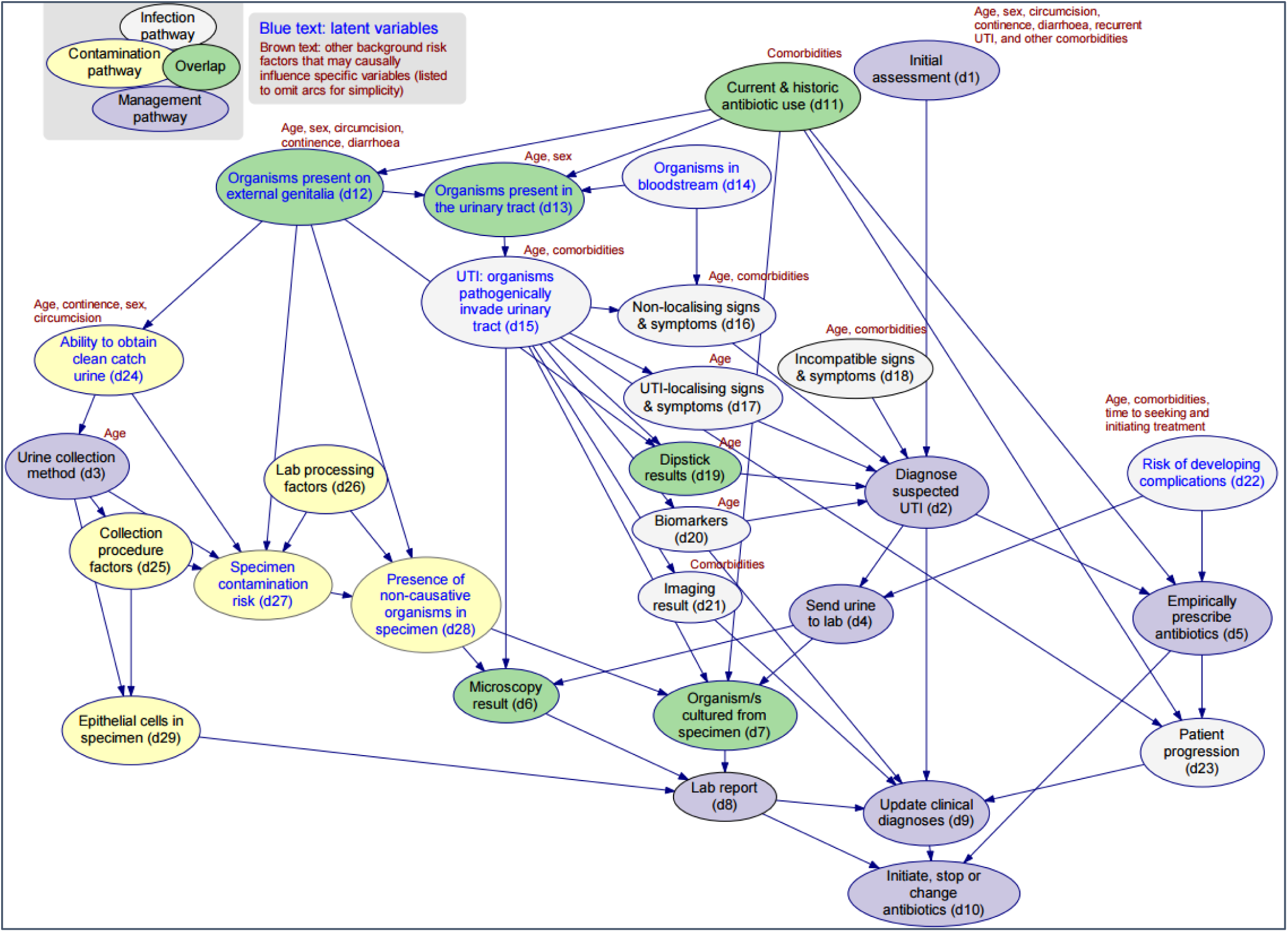
The Expert DAG v11.1, as provided in the main manuscript (Figure 1).

**Table C1.**
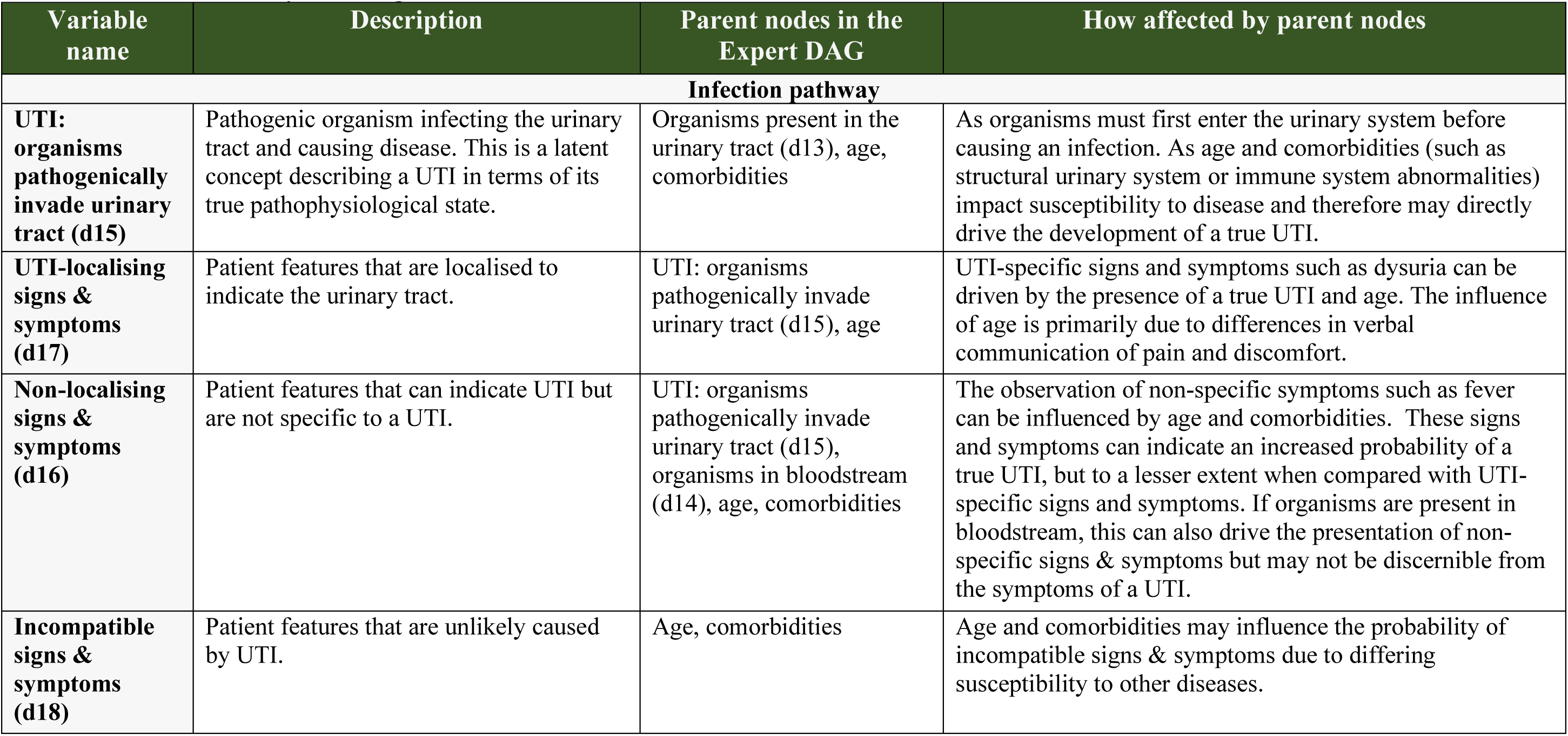
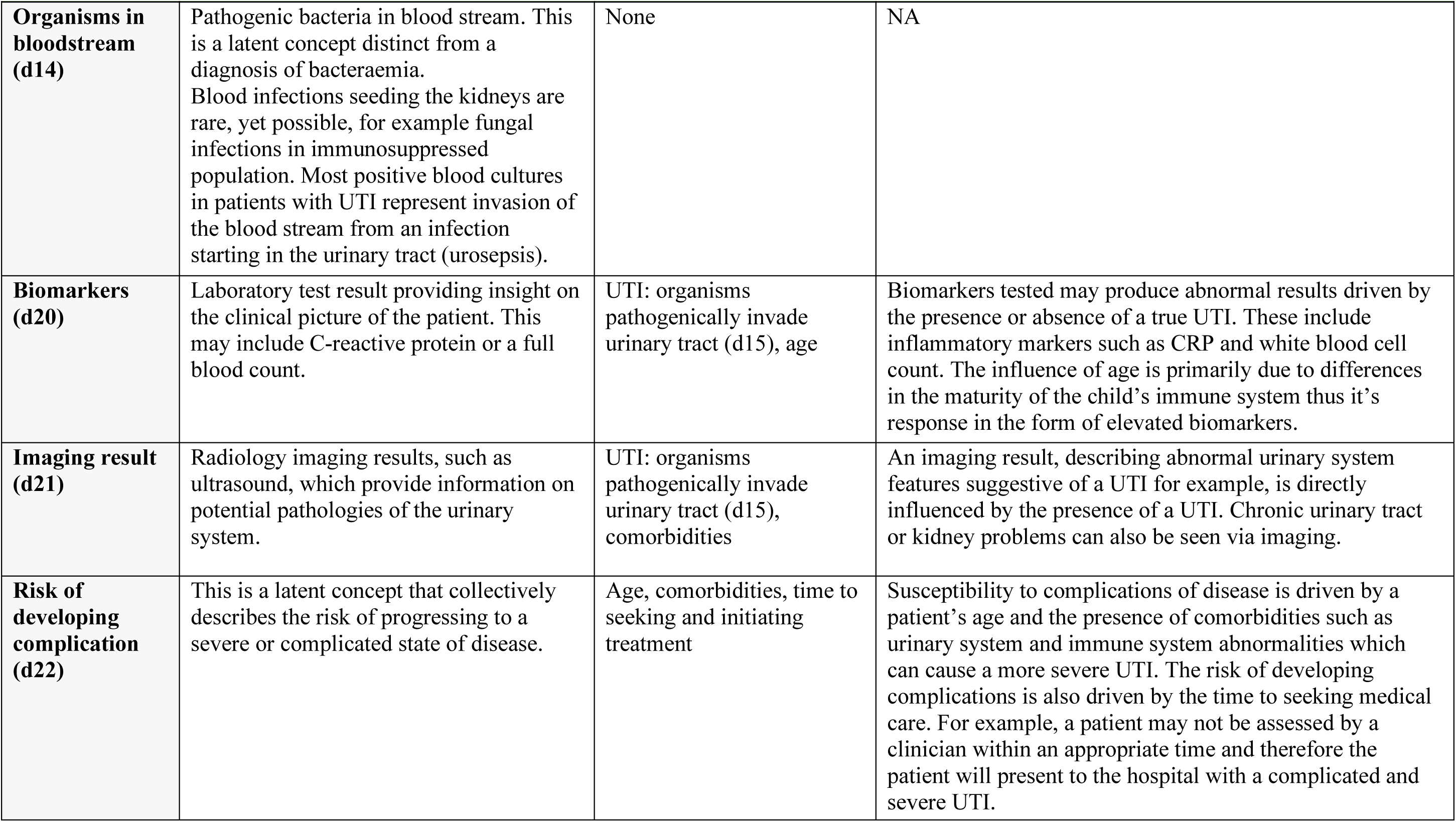
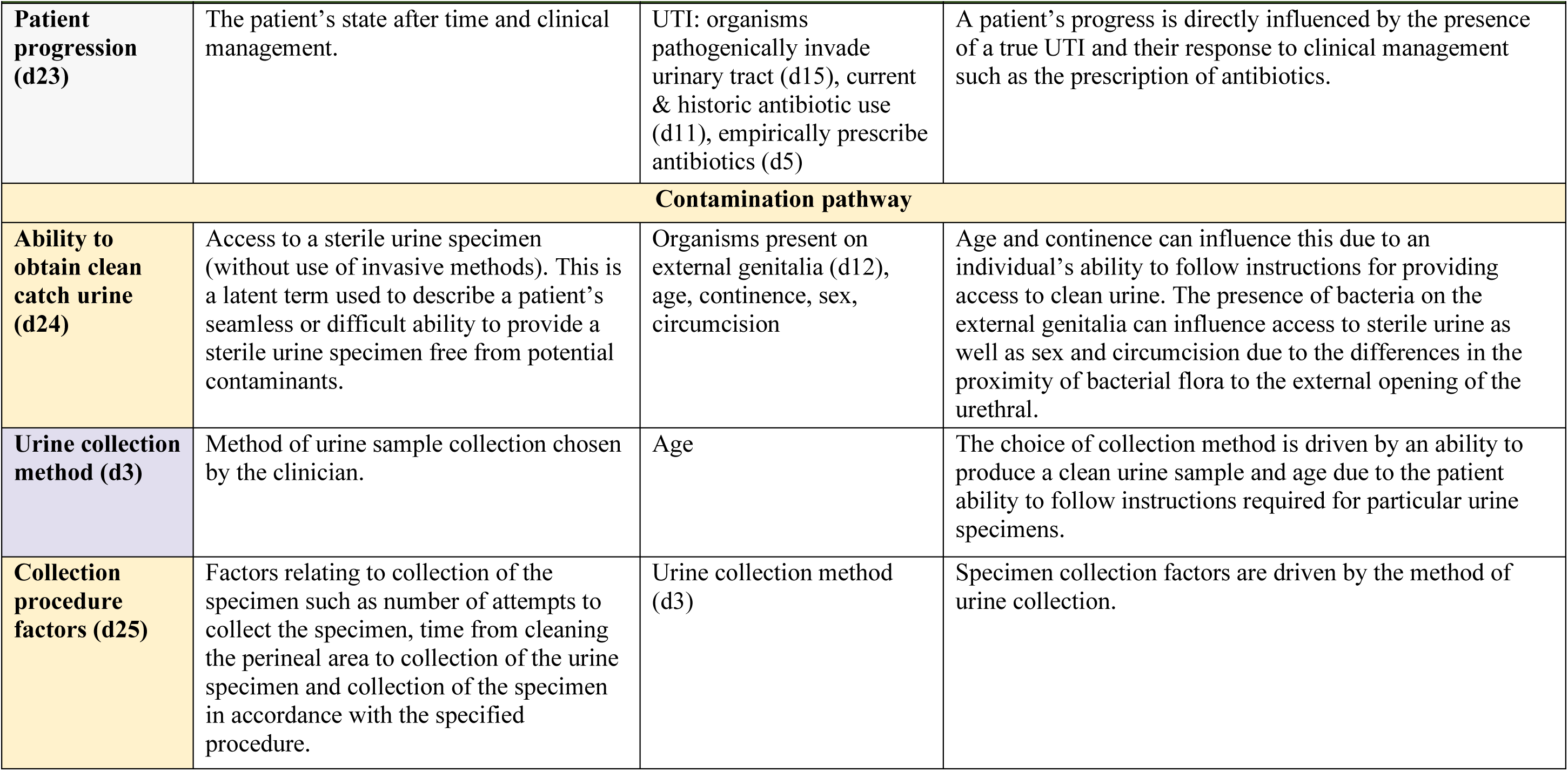
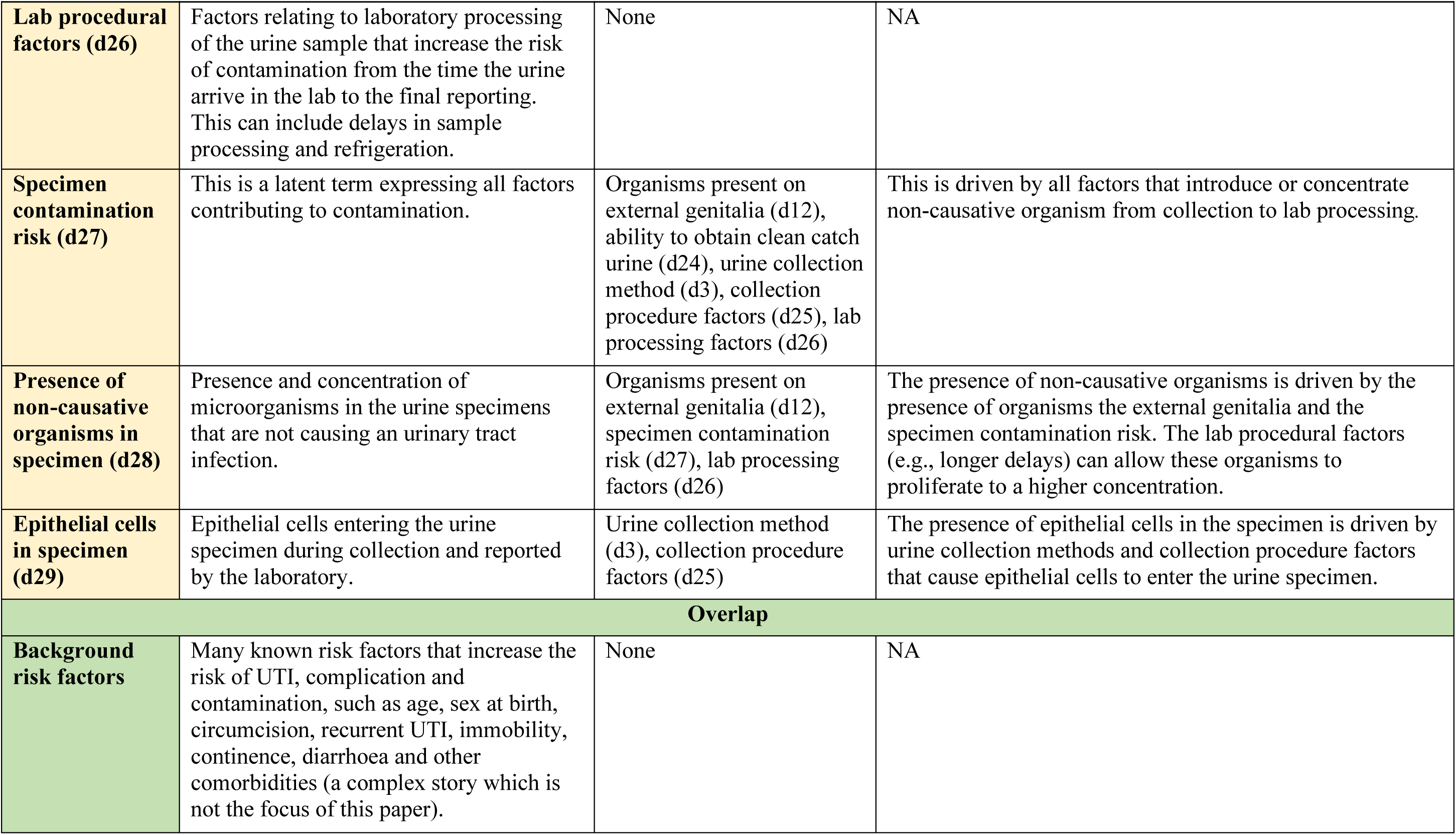
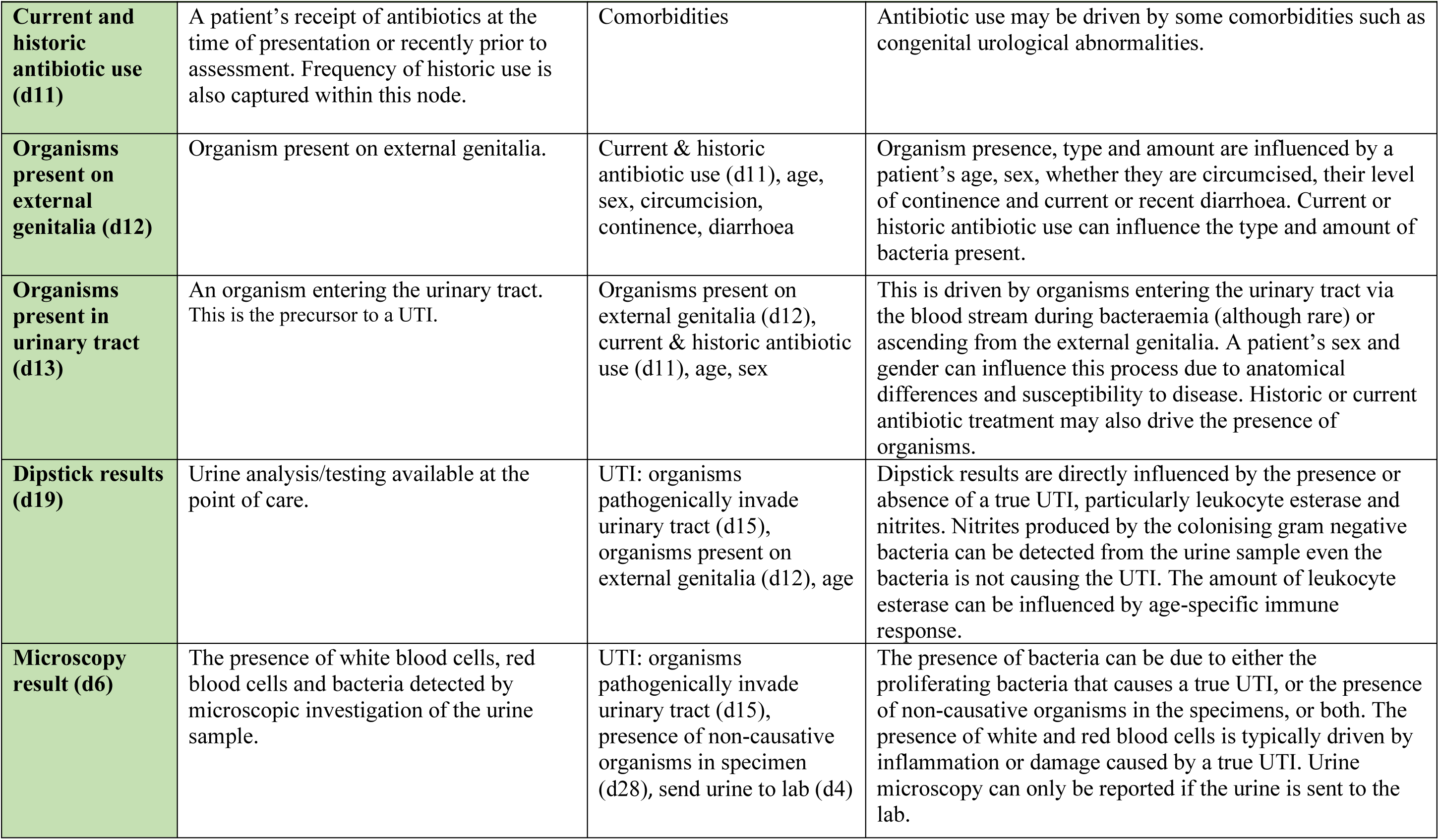
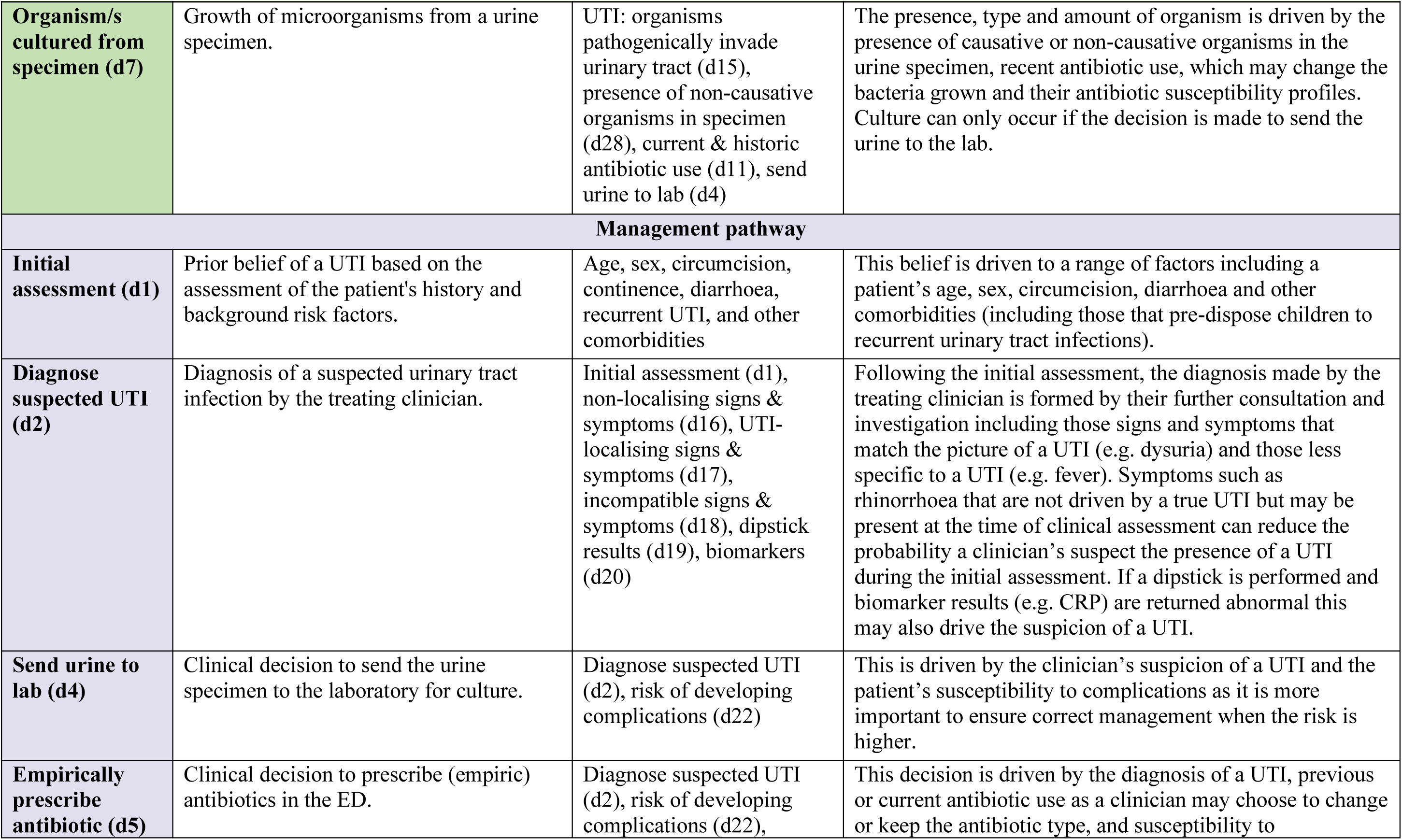
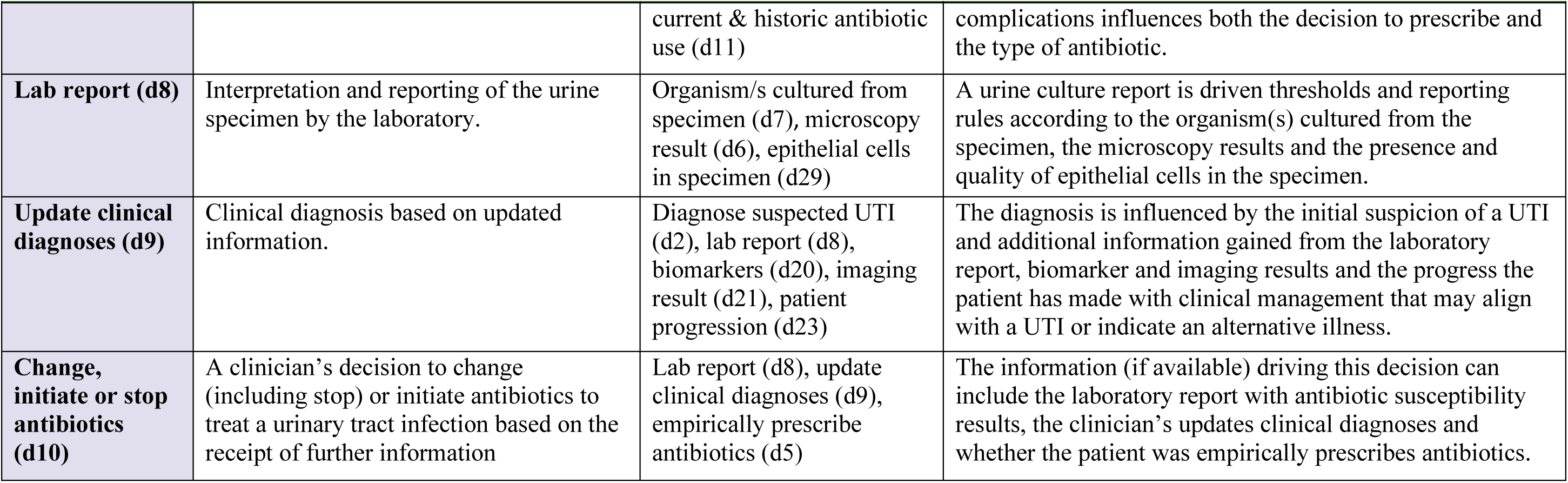
Variable dictionary of the Expert DAG v11.1.

## Additional file 4: List of changes when converting the Expert DAG to the Applied BN

In this document we summarised the major considerations when converting the Expert DAG to the Applied BN. Conversion of the Expert DAG took into consideration: how a particular variable is relevant to the applied BN’s purpose; how it could be matched to available data; and how it could help simplify parameterisation or computational workload. This frequently involved simplifications by removing and merging variables, as well expansions by splitting and adding variables. Figure D1 illustrates an example list of decisions made on whether to keep a DAG variable in the BN. We also provided a full list of changes occurred during the conversion from the Expert DAG v11.1 to the Applied BN v2.2 (Table D1).

**Figure D1.**
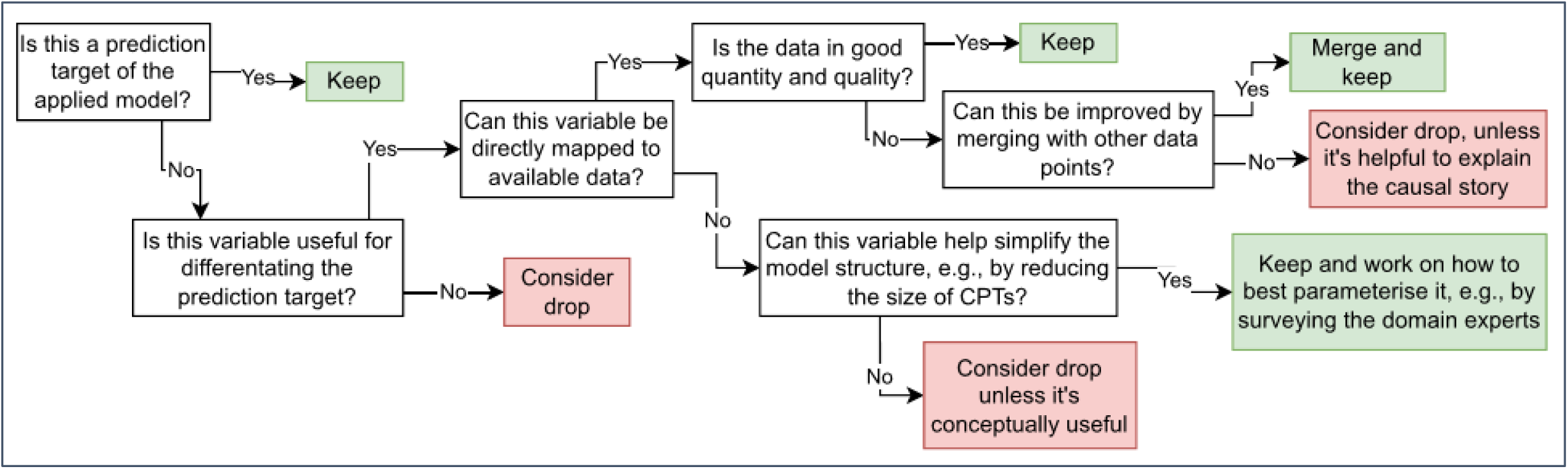
An example procedure of deciding on whether to keep a DAG variable in the Applied BN.

**Table D1.** Structural changes occurred during our conversion from the Expert DAG v11.1 to the Applied BN v2.2 and their underlying rationale.

| **Changes** | **Expert DAG** | **Applied BN** |
| --- | --- | --- |
| Route of UTI and causative pathogens | Organisms present on external genitalia (d12) and organisms in bloodstream (d14) are two possible routes that can lead to organisms present in the urinary tract (d13) and subsequently UTI (d15). | Acquiring UTI from bloodstream (d14) is rare, thus was dropped for simplification. Grouping d12 and d13 together as local colonisation, which approximates a broader area where presence of organism is relevant (possible to cause UTI). Splitting a general organism concept into three specific organism groups (b7-9), which are the primary predication targets of the model. |
| Ability to obtain clean catch urine | Explicitly described as a concept that can be influenced by age and sex etc. (d24), and subsequently drives urine collection methods (d3), contamination risk (d27). | This concept node is removed to simplify the parameterisation need, instead, relevant background factors directly go into urine collection methods (b13) and contamination risk (b15) to maintain the associations. |
| Presence of non-causative organism in specimen | An intermediate step that explicitly describes how (non-causative) colonising organisms may be present in specimen thus isolated (d28). | This intermediate node was removed for simplicity, as a result, the local colonisations (b7-b9) directly influence the culture results (b17-b19). |
| Collection, lab procedure factors | Described using d25-26 for completeness of the causal story. | Dropped as considered not highly influential by the domain experts, as well as with insufficient data for investigation. |
| Background risk factors | Simplified as brown text. | Key ones explicitly included as variables/nodes: age group (b1), UTI-relevant comorbidity (b3), sex (b2) and diarrhoea (b4). |
| Culture results | Summarised as a single node (d7) | Organism group specific nodes b17-19 |
| Symptoms and signs | Single node was used to summarise a group of signs and symptoms, including those are UTI localising (d17), non-localising (d16), and incompatible with UTI (d18). | Specific signs and symptoms are described using separate nodes/variables with certain interactions among the signs and symptoms (b26-37). Looking for differential effects of different signs and symptoms. Observations can be made from the presenting patients, entered as input variables when applying the model. |
| Dipstick | Included as a summary node (d19) | Expanded for differential effects based on data availability (b20-21). |
| Biomarkers | Included as a summary node (d20) | Expanded for differential effects based on data availability (b23-25). |
| Microscopy result | Included as a summary node (d6) | Expanded for differential effects based on data availability (b16, b22). |
| Initial assessment, diagnosis of suspected UTI | d1-2, clinical diagnosis based on evidence available to treating doctor at the point of care. | Suspected UTI was an inclusion criteria for the PEA study cohort, thus these variables were removed. As a result, we are not able to explore how such clinical diagnosis was made. |
| Send urine to lab | d4 | This was an inclusion criteria for the PEA study cohort of suspected UTI, thus this variable is removed. As a result, we are not able to explore how the urine test decision was made. |
| Lab report | d8, included to summarise results on microscopy results (d6), epithelial cells (d29) and culture results (d7). | Dropped as the microscopic analysis (b16, b22), epithelial cells (b14), and culture results (b17-19) were explicitly included. |
| Propensity of UTI Progression | Risk of developing complications (d22) | This variable was first divided into two concepts: susceptibility to UTI progression (b5) and current UTI severity (b11). After further workshop discussion, b5 and b11 were merged as one variable (for structure simplicity) and renamed as current clinical phenotype (b11). |
| Patient outcome of the current episode | Described using patient progression (d23), update clinical diagnoses (d9), and initiate, stop or change antibiotics (d10). | All dropped as they are out of the scope of the current application. |

## Additional file 5: The Applied BN and dictionary

In this document we provided the detailed structure of the Applied BN v2.2 include a high-level structure (Figure E1) and 4 submodels (Figures E2-E5) and the variable dictionary of this model (Table E1).

**Figure E1.**
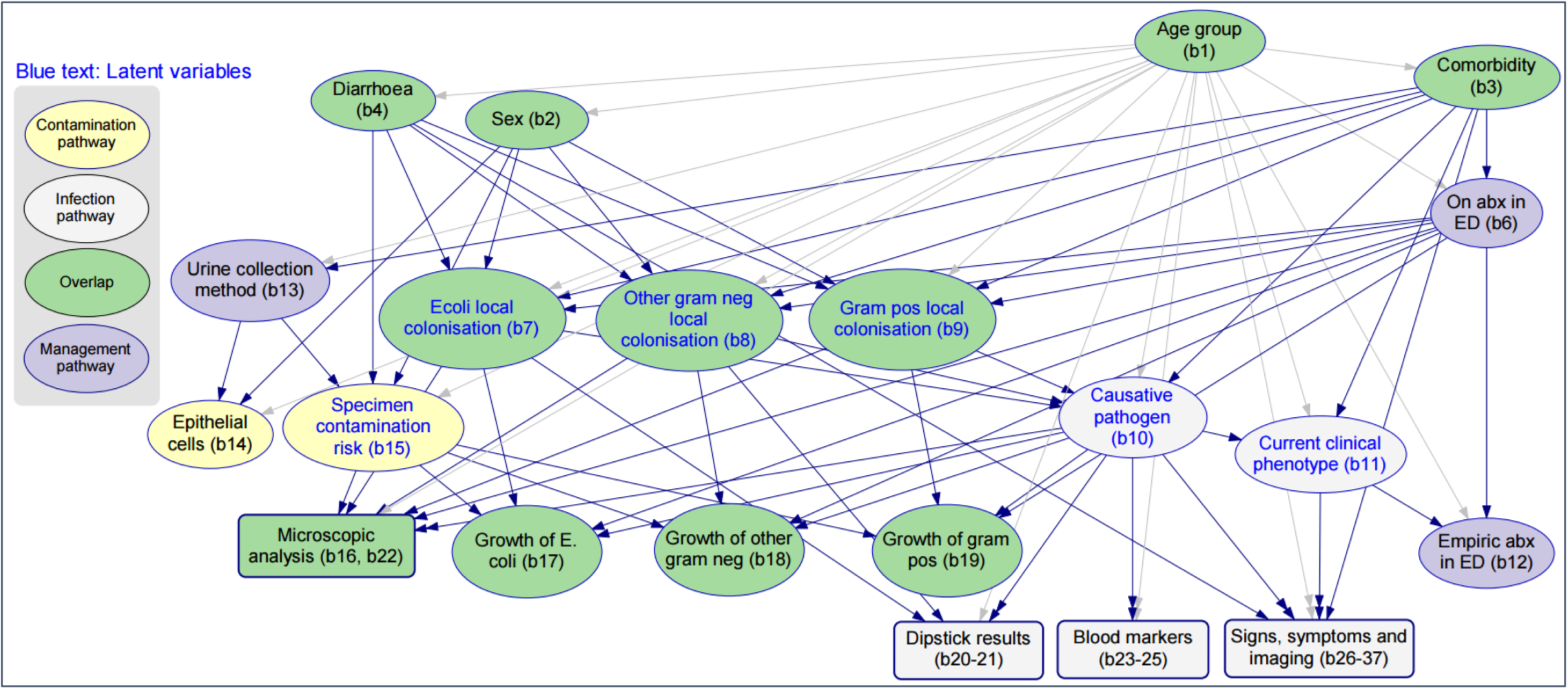
The high-level structure of the Applied BN v2.2, as provided in the main manuscript (Figure 2, bottom panel).

**Figure E2.**
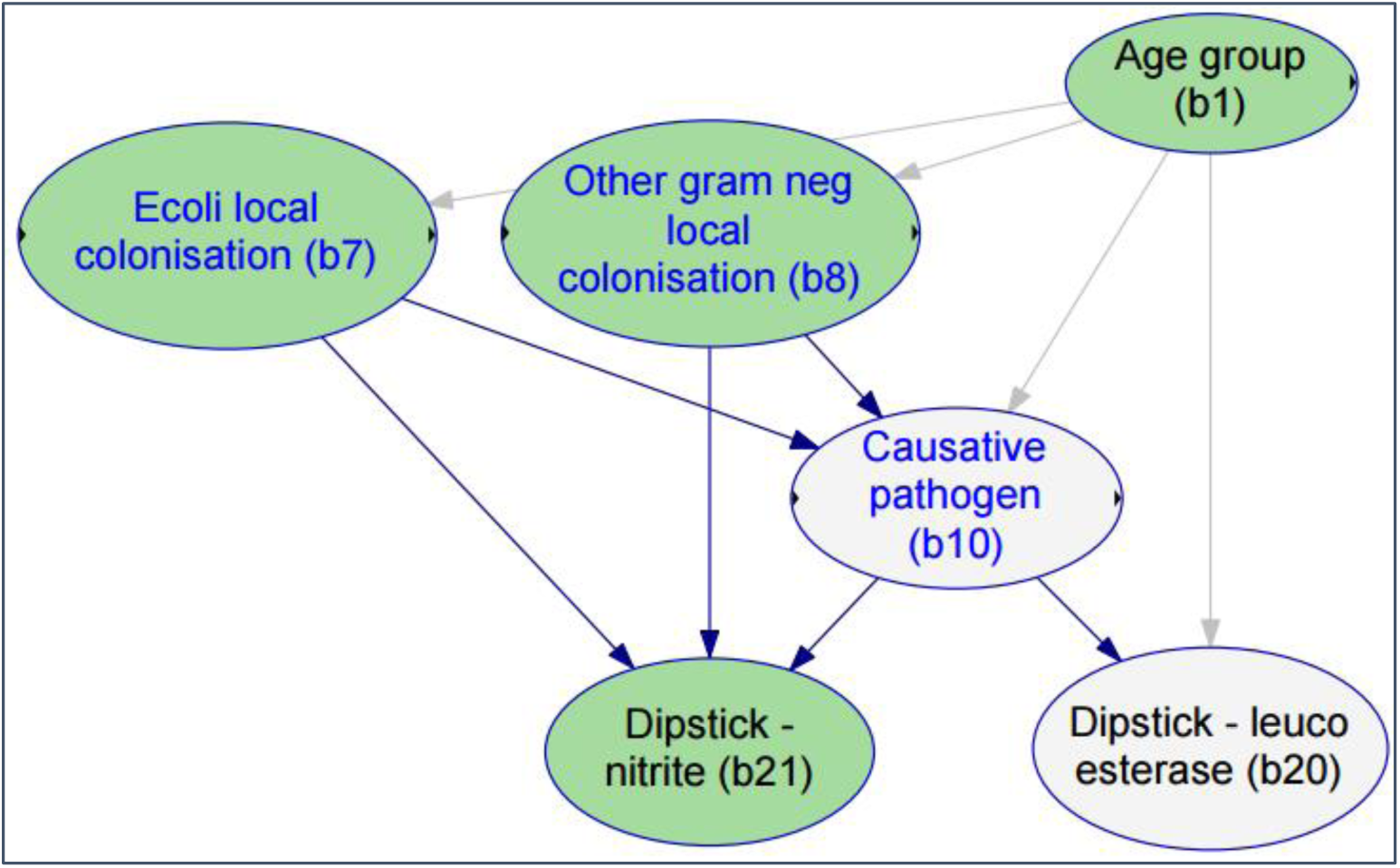
The local structure of dipstick results submodel (b20-21) with external connections (b1, b7-8, b10).

**Figure E3.**
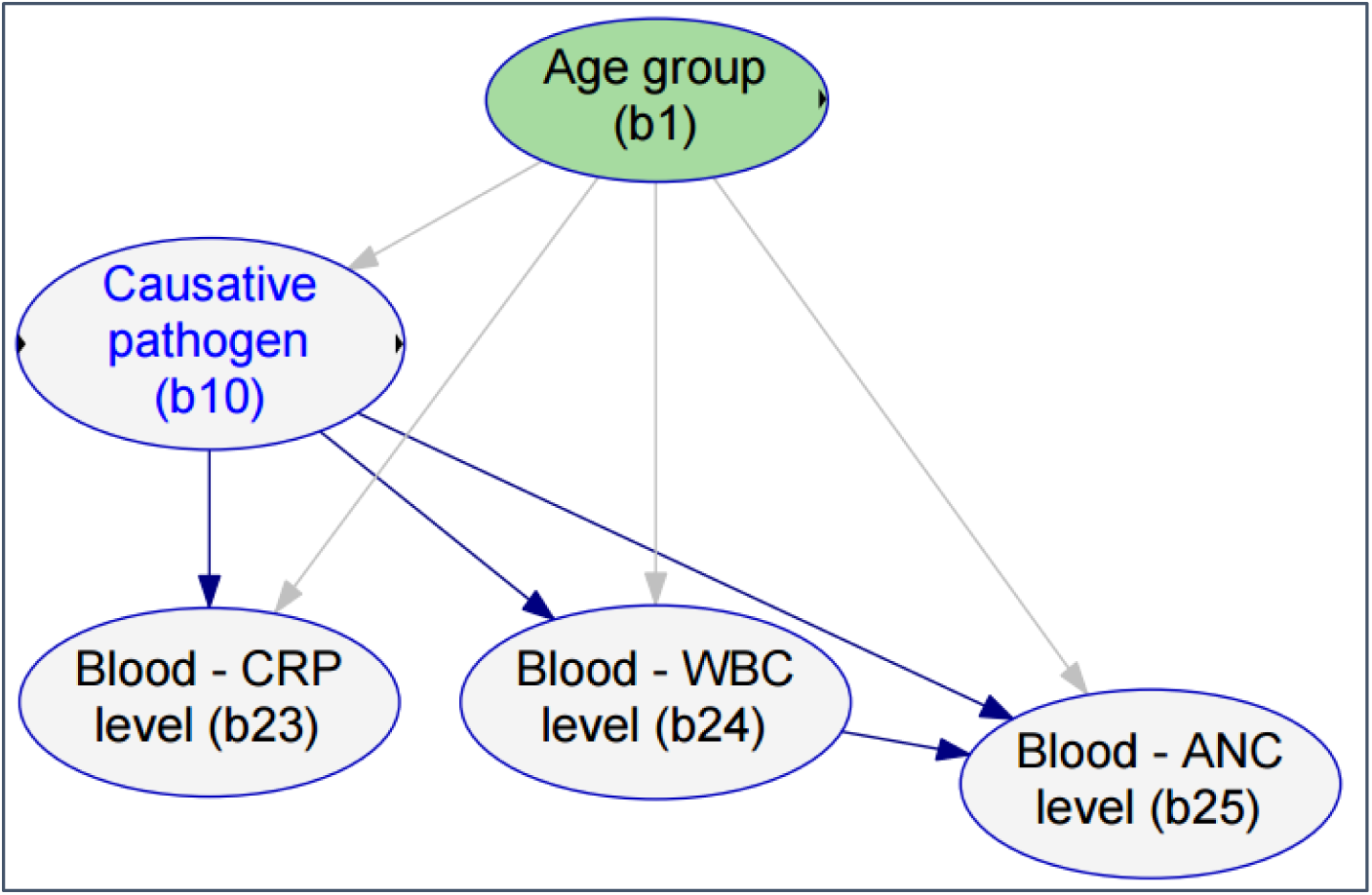
The local structure of microscopic analy (b23-25) with external connections (b1, b10).

**Figure E4.**
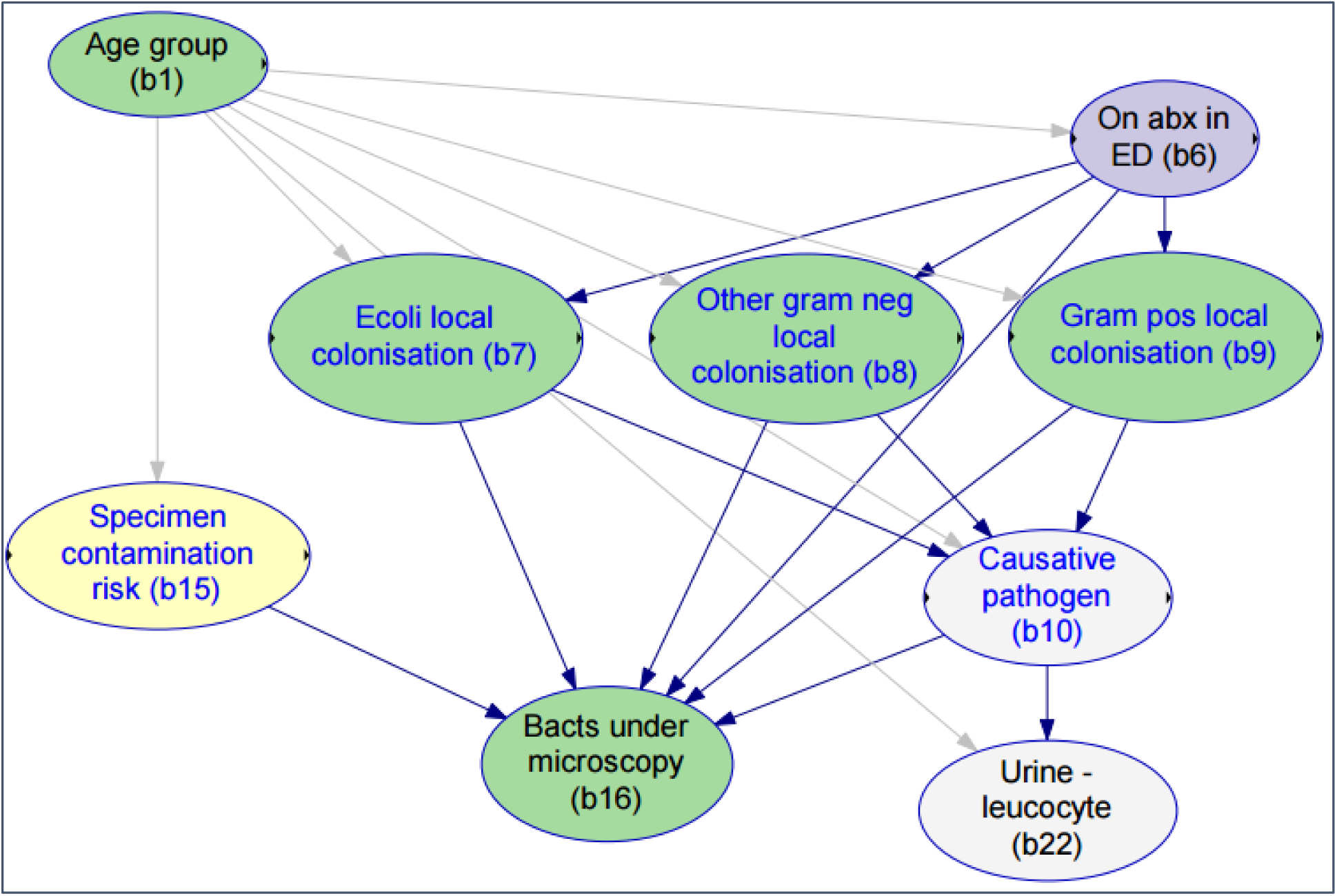
The local structure of microscopic analysis submodel (b16, b22) with external connections (b1, b6, b7-10, b15).

**Figure E5.**
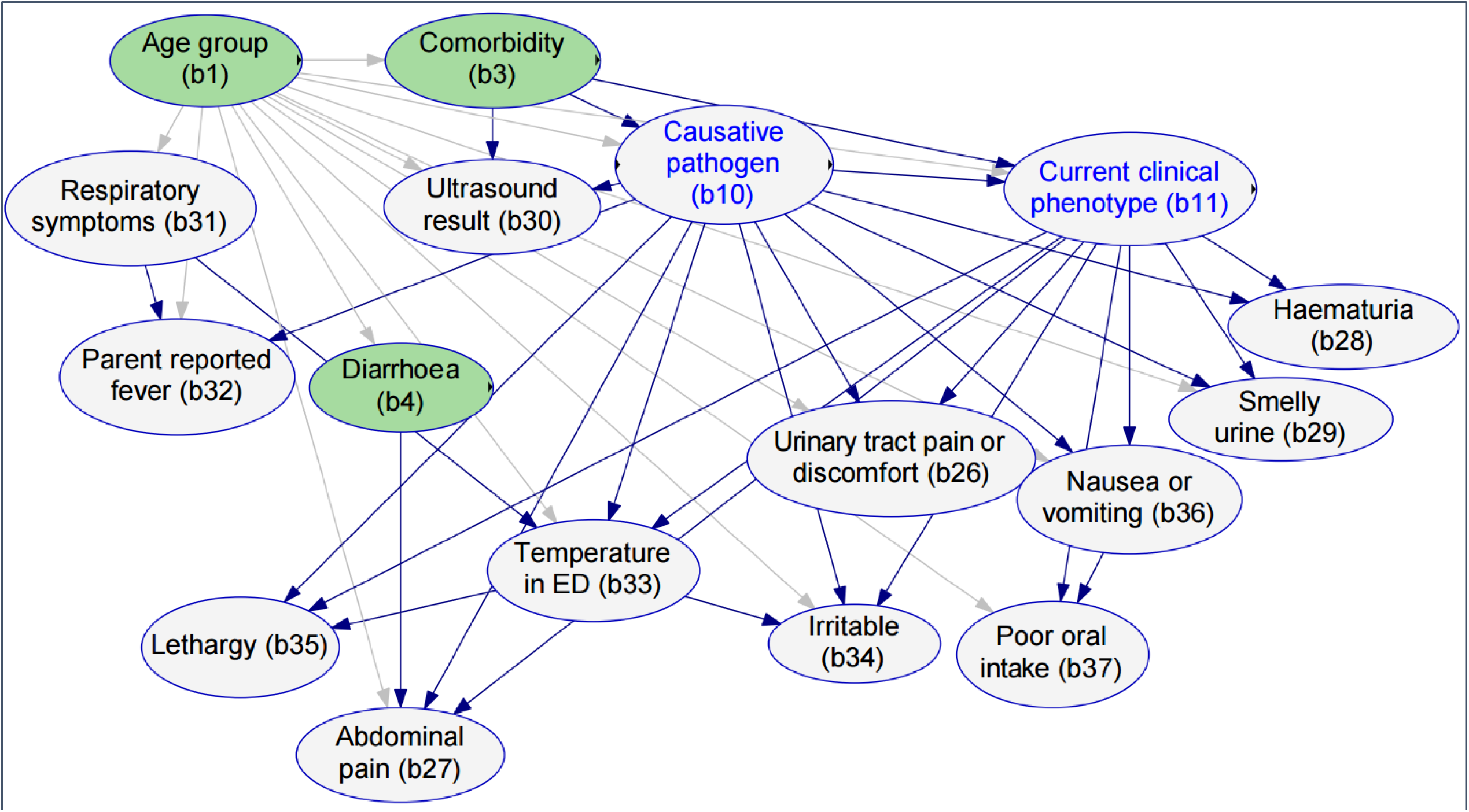
The local structure of signs, symptoms and imaging (b26-b37) with external connections (b1, b3-4, b10, b11). Of note, although the history of diarrhoea (b4) can be a symptom caused by UTI, we modelled it as an important possible cause of UTI therefore excluded from the signs, symptoms and imaging submodel. However, the presence of diarrhoea may lead to reported abdominal pain, thus shown here as an external connection.

**Table E1.** The variable dictionary of the Applied BN v2.1, and how the variables correspond to the Expert DAG v11.1.

| vid | Variable label | Definition | States | Parents | Corresponding node/s in the Expert DAG v11 | Status |
| --- | --- | --- | --- | --- | --- | --- |
| b1 | **Age group** | Age group. All children under 13yo as an inclusion criteria for the PEA Study. | *LessThan6Mon, Btw6MonAnd2Yr, Btw2And5Yr, Above5Yr* | None | Simplified as brown text | Observable |
| b2 | **Sex** | Sex at birth. | *Female, Male* | Age group | Simplified as brown text | Observable |
| b3 | **Comorbidity** | History of UTI or a history of urinary tract problems, either recorded within their medical notes or reported by their carers in the PEA study survey. | *Reported, Unknown* | Age group | Simplified as brown text | Observable |
| b4 | **Diarrhoea** | History of diarrhoea. Although this can be a symptom caused by UTI, we modelled it as an important possible cause of UTI rather than caused by. | *Yes, No* | Age group | Simplified as brown text | Observable |
| b6 | **On abx in ED** | On antibiotics upon presentation to ED, if yes, the specified antibiotic was classified as narrow ($\leq 3$) or broader ($> 3$) according to published Antibiotic Spectrum Index (Gerber et al., 2017)<sup>1</sup>. | *Narrow, Broader, No* | Age group, comorbidity | d11 | Observable |
| b7 | **E.coli local colonisation** | The level of colonisation of the perineum/ external genitalia by E. coli., this is assumed to predispose children to E. coli UTI. | *High, Low* | Age group, sex, comorbidity, diarrhoea, on abx in ED | d12, d13 | Latent |
| b8 | **Other gram neg local colonisation** | The level of colonisation of the perineum/ external genitalia by non-E. coli gram negative bacteria, this is assumed to predispose children to other gram negative UTI. | *High, Low* | Age group, sex, comorbidity, diarrhoea, on abx in ED | d12, d13 | Latent |
| b9 | **Gram pos local colonisation** | The level of colonisation of the perineum/ external genitalia by gram positive bacteria, this is assumed to predispose children to gram positive UTI. | *High, Low* | Age group, sex, comorbidity, diarrhoea, on abx in ED | d12, d13 | Latent |
| b10 | **Causative pathogen** | The pathogenic organism infecting the urinary tract and causing disease. | *EColi, OtherGramNeg, GramPos, None* | Age group, comorbidity, E.coli local colonisation, other gram neg local colonisation, gram pos local colonisation | d15 | Latent |
| b11 | **Current clinical phenotype** | This was introduced as a summary node of patient presentation phenotypes based on signs and symptoms relevant to UTI. | *Type 1, Type 2, Type 3* | Age group, comorbidity, causative pathogen | d22 | Latent<sup>2</sup> |
| b12 | **Empiric abx in ED** | Empiric antibiotic prescription received during the ED presentation. This is an inclusion criteria for the PEA Study. | *Narrow, Broader* | On abx in ED, current clinical phenotype, age group | d5 | Observable |
| b13 | **Urine collection method** | Method of urine sample collection chosen by the clinician. | *CleanCatch, SupraAsp, Catheter* | Age group, comorbidity | d3 | Observable |
| b14 | **Epithelial cells** | Assessment of epithelial cells via microscopic or automated analysis. | *Moderate, Low* | Age group, sex, urine collection method | d29 | Observable |
| b15 | **Specimen contamination risk** | This latent concept refers to the risk of a non-causative organism/s entering the urine specimen during the specimen collection process. | *High, Low* | Age group, sex, diarrhea, urine collection method | d27 | Latent |
| b16 | **Bacts under microscopy** | Assessment of bacteria via microscopic analysis. | *Many, Moderate, Few, NotSeen* | Causative pathogen, on abx in ED, specimen contamination risk, E.coli local colonisation, other gram neg local colonisation, gram pos local colonisation | d6 | Observable |
| b17 | **Growth of E.coli** | The isolation of E.coli from the urine specimen. | *Positive, Negative* | E.coli local colonisation, specimen contamination risk, on abx in ED, causative pathogen | d7 | Observable |
| b18 | **Growth of other gram neg** | The isolation of non-E.coli gram negative bacteria from the urine specimen. | *Positive, Negative* | Other gram neg local colonisation, specimen contamination risk, on abx in ED, causative pathogen | d7 | Observable |
| b18 | **Growth of gram pos** | The isolation of gram positive bacteria from the urine specimen. | *Positive, Negative* | Gram pos local colonisation, specimen contamination risk, on abx in ED, causative pathogen | d7 | Observable |
| b20 | **Dipstick - leuco esterase** | Leukocyte esterase detected on urine dipstick. | *High, Moderate, Low, NotDetected* | Causative pathogen, age group | d19 | Observable |
| b21 | **Dipstick - nitrite** | Nitrite detected on the urine dipstick. | *Detected, NotDetected* | Causative pathogen, E.coli local colonisation, other gram neg local colonisation | d19 | Observable |
| b22 | **Urine - leucocyte** | The assessment of leucocytes level in the urine specimen via microscopic analysis. | *High, Moderate, Low* | Causative pathogen, age group | d6 | Observable |
| b23 | **Blood - WBC level** | Leukocyte counts in blood. | *Above18, Btw10And18, Below10, NotDone* | Causative pathogen, age group | d20 | Observable |
| b24 | **Blood - CRP level** | C-Reactive protein in blood. | *Above70, Btw15And70, Below15, NotDone* | Causative pathogen, age group | d20 | Observable |
| b25 | **Blood - ANC level** | Absolute neutrophil counts in blood. | *Above15, Btw8And15, Below8, NotDone* | Causative pathogen, age group, blood - WBC level | d20 | Observable |
| b26 | **Urinary tract pain or discomfort** | Genital pain (or discomfort), dysuria, and urinary tract relevant discomfort based on free text in medical notes recorded during ED presentation. | *Yes, Unknown* | Causative pathogen, current clinical phenotype, age group | d17 | Observable |
| b27 | **Abdominal pain** | Abdominal pain in medical notes recorded during ED presentation. | *Yes, Unknown* | Causative pathogen, current clinical phenotype, age group, diarrhea | d16, d17 | Observable |
| b28 | **Haematuria** | Haematuria in medical notes recorded during ED presentation. | *Yes, Unknown* | Causative pathogen, current clinical phenotype | d17 | Observable |
| b29 | **Smelly urine** | Foul smelling urine in medical notes recorded during ED presentation. | *Yes, Unknown* | Causative pathogen, current clinical phenotype, age group | d17 | Observable |
| b30 | **Ultrasound result** | Result of ultrasound investigation. Abnormal ultrasound result was defined if there were features suggestive of pyelonephritis, renal abscess, cystitis or other UTI evidence. | *Abnormal, unknown, NotDone* | Age group, causative pathogen, comorbidity | d21 | Observable |
| b31 | **Respiratory symptoms** | Respiratory symptoms in medical notes recorded during ED presentation. | *Yes, No* | Age group | d18 | Observable |
| b32 | **Parent reported fever** | Parent reported fever in medical notes recorded during ED presentation. | *Yes, No* | Causative pathogen, age group, respiratory symptoms | d16 | Observable |
| b33 | **Temperature in ED** | Temperature (degrees celsius) recorded in ED. | *Abv385, Btw375and385, Btw365and375, Below365* | Causative pathogen, current clinical phenotype, age group, respiratory symptoms | d16 | Observable |
| b34 | **Irritable** | Irritability in medical notes recorded during ED presentation. | *Yes, No* | Current clinical phenotype, age group, causative pathogen, temperature in ED | d16 | Observable |
| b35 | **Lethargy** | Lethargy in medical notes recorded during ED presentation. | *Yes, No* | Causative pathogen, current clinical phenotype, temperature in ED | d16 | Observable |
| b36 | **Nausea or vomiting** | Nausea and/or vomiting in medical notes recorded during ED presentation. | *Yes, No* | Causative pathogen, current clinical phenotype, age group | d16 | Observable |
| b37 | **Poor oral intake** | Poor oral intake in medical notes recorded during ED presentation. | *Yes, No* | Current clinical phenotype, age group, nausea or vomiting | d16 | Observable |

<sup>1</sup>Narrow: Amoxicilin, Trimethoprim, Benzylpenicillin, Cefalexin, Cefazolin, Erythromicin. Broader: Amoxicillin + Clavulanic acid, Trimethoprim + Sulfamethoxazole, Co-trimoxazole, Amikacin, Cefepime, Cefotaxime, Ceftazidime, Ceftriaxone, Ciprofloxacin, Colistin, Ertapenem, Gentamicin, Meropenem, Moxifloxacin, Nitrofurantoin, Norfloxacin, Piperacillin + Tazobactam, Tazocin, Tobramycin, Vancomycin
<sup>2</sup>This variable is latent, but was treated uniquely in order to provide a definition of current clinical phenotype that is independent of other latent factors in the model. In particular, a separate clustering was performed (using the EM algorithm) on the signs and symptoms, resulting in a grouping into three types, simply called “Type 1”, “Type 2” and “Type 3”, “Type 1” being systemic signs and symptoms predominant but mild urinary tract localising symptoms, “Type 2” being urinary tract localising symptoms predominant, and “Type 3” being abdominal pain predominant with minor other symptoms.

## Additional file 6: Parameterisation survey responses

In this document we provided a summary of survey responses used to inform the BN parameters.

### Q1. Risk of specimen contamination

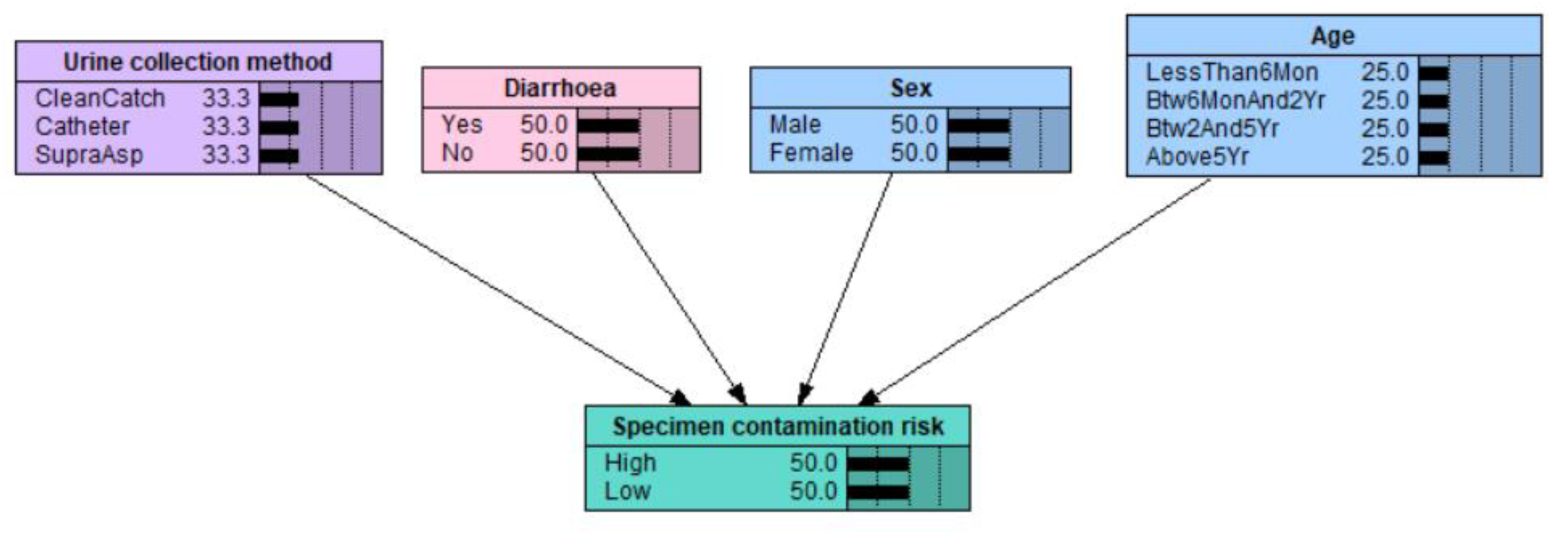

Consider the risk of a non-causative organism/s entering the urine specimen during the specimen collection process. In the model (as shown in the above figure), the **risk of specimen contamination** is influenced by **age**, **sex**, presence of **diarrhoea**, and **urine collection method**. Assuming *the same* colonisation status of each child’s perineum/ external genitalia (i.e., type and density of organisms), how do the following factors increase or decrease the risk of specimen contamination from the baseline (as specified below)? E.g., x0.3, x2, x10, etc. Mean and standard deviation based on survey responses from 8 experts.

1a. **Age** and **sex**, assuming **clean catch** as the method of specimen collection.

| Age | Male, mean (sd) | Female, mean (sd) |
| --- | --- | --- |
| >=5yo | 1 (baseline) | 1.8 (1.0) |
| 2 to 5yo | 2.2 (0.7) | 3.6 (2.0) |
| 6mon to 2yo | 3.3 (1.4) | 5.2 (3.1) |
| <6mon | 4.2 (2.1) | 6.6 (4.0) |

1b. Presence of **diarrhoea**, assuming **clean catch** as the method of specimen collection.

|  |  |
| --- | --- |
| Absence | 1 (baseline), mean (sd) |
| Presence | 4.1 (1.6) |

1c. **Urine collection method**

|  |  |
| --- | --- |
| Supra aspirate | 1 (baseline), mean (sd) |
| Catheter | 2.3 (1.2) |
| Clean catch | 6.8 (5.4) |

**Collated comments**

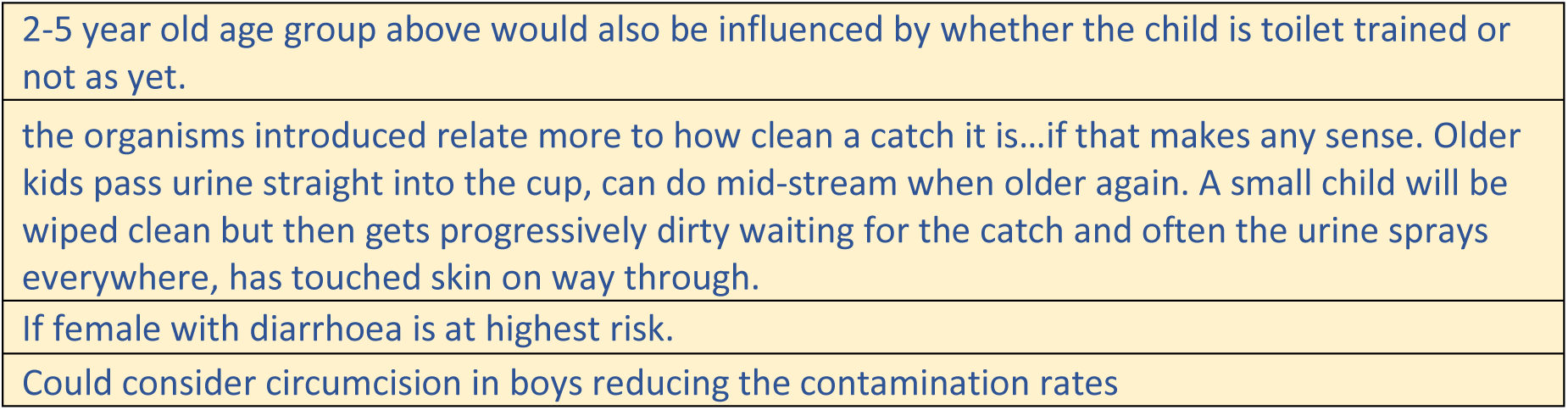

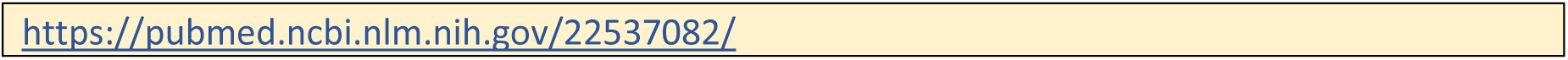

### Q2. Propensity to UTI progression

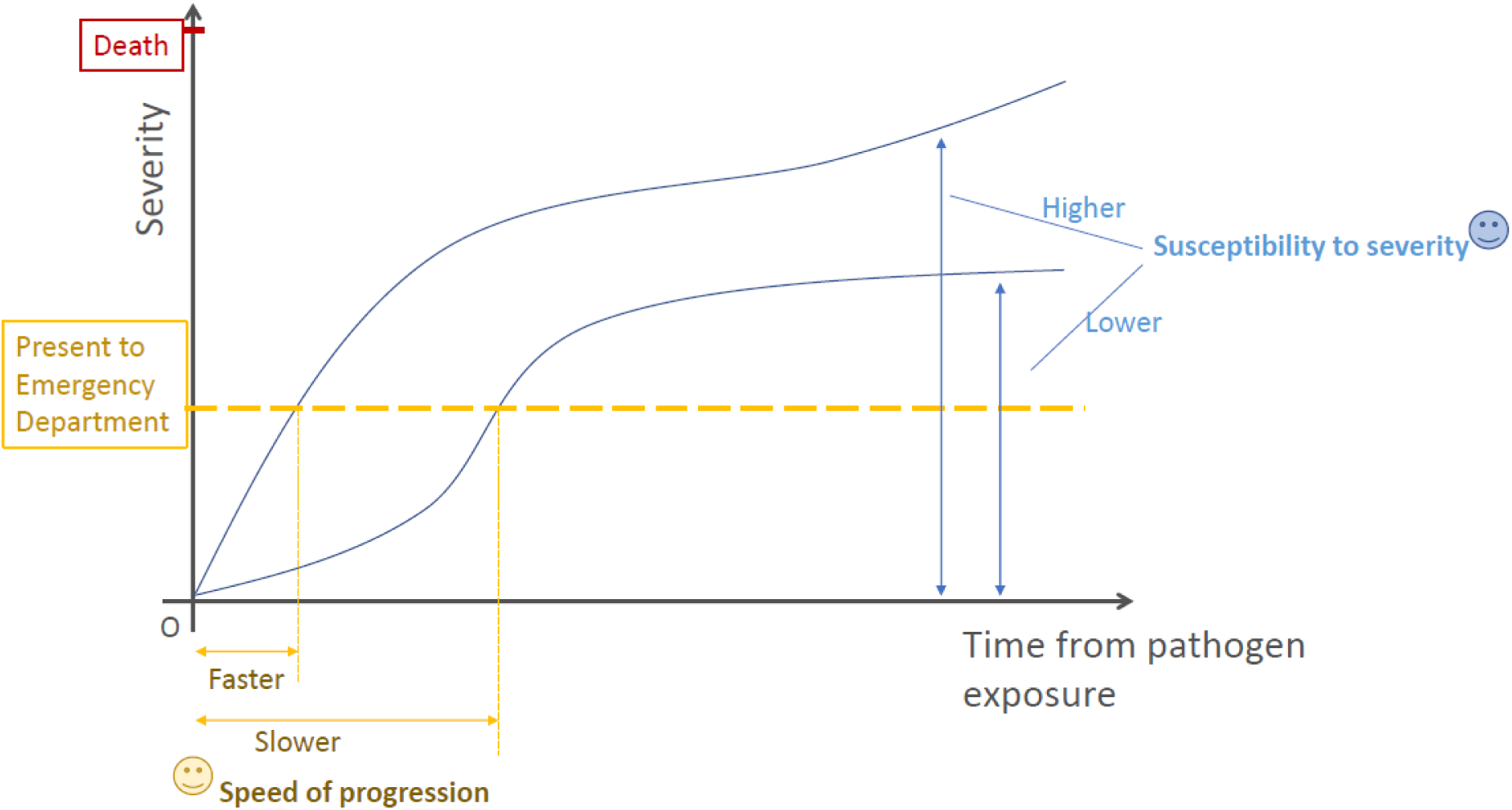

In the model, both the **speed of progression** and **susceptibility to severity** may be influenced by **age** and **UTI-relevant comorbidity** (such as VUR/anatomical abnormalities of the urinary tract). We now ask a series of questions on these two concept variables. Please provide your min, max, and best guess estimates for each question. Please note that the “min/max” should be plausible lower or upper values, e.g., 95th percentiles, not the extreme recordable value. Mean and standard deviation based on survey responses from 8 experts.

#### 2a. Speed of progression

**Age**

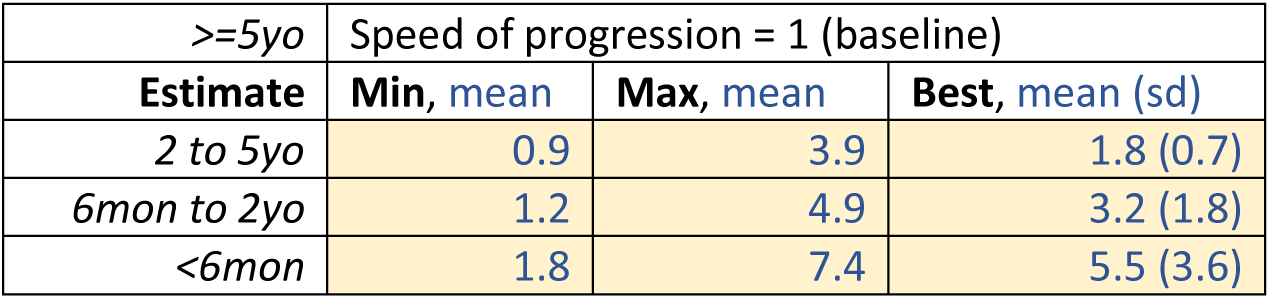

**UTI-relevant comorbidity**

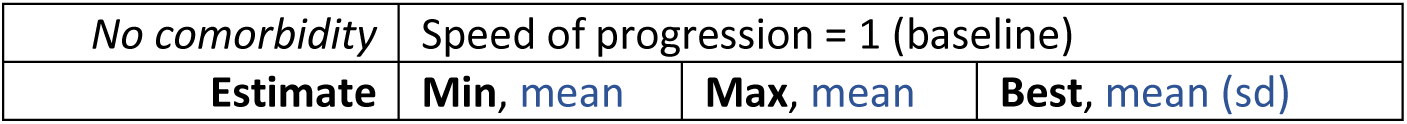

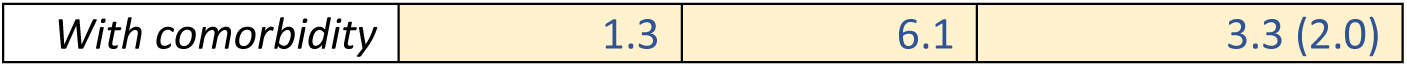

#### 2b. Susceptibility to severity

**Age**

| *>=5yo* | Susceptibility to severity = 1 (baseline) |  |  |
| --- | --- | --- | --- |
| Estimate | Min, mean | Max, mean | Best, mean (sd) |
| *2 to 5yo* | 1.0 | 4.1 | 1.7 (0.8) |
| *6mon to 2yo* | 1.2 | 4.8 | 3.0 (2.0) |
| *<6mon* | 1.7 | 6.3 | 5.1 (3.9) |

**UTI-relevant comorbidity**

| *No comorbidity* | Susceptibility to severity = 1 (baseline) |  |  |
| --- | --- | --- | --- |
| Estimate | Min, mean | Max, mean | Best, mean (sd) |
| *With comorbidity* | 1.4 | 6.3 | 3.6 (1.8) |

Collated comments

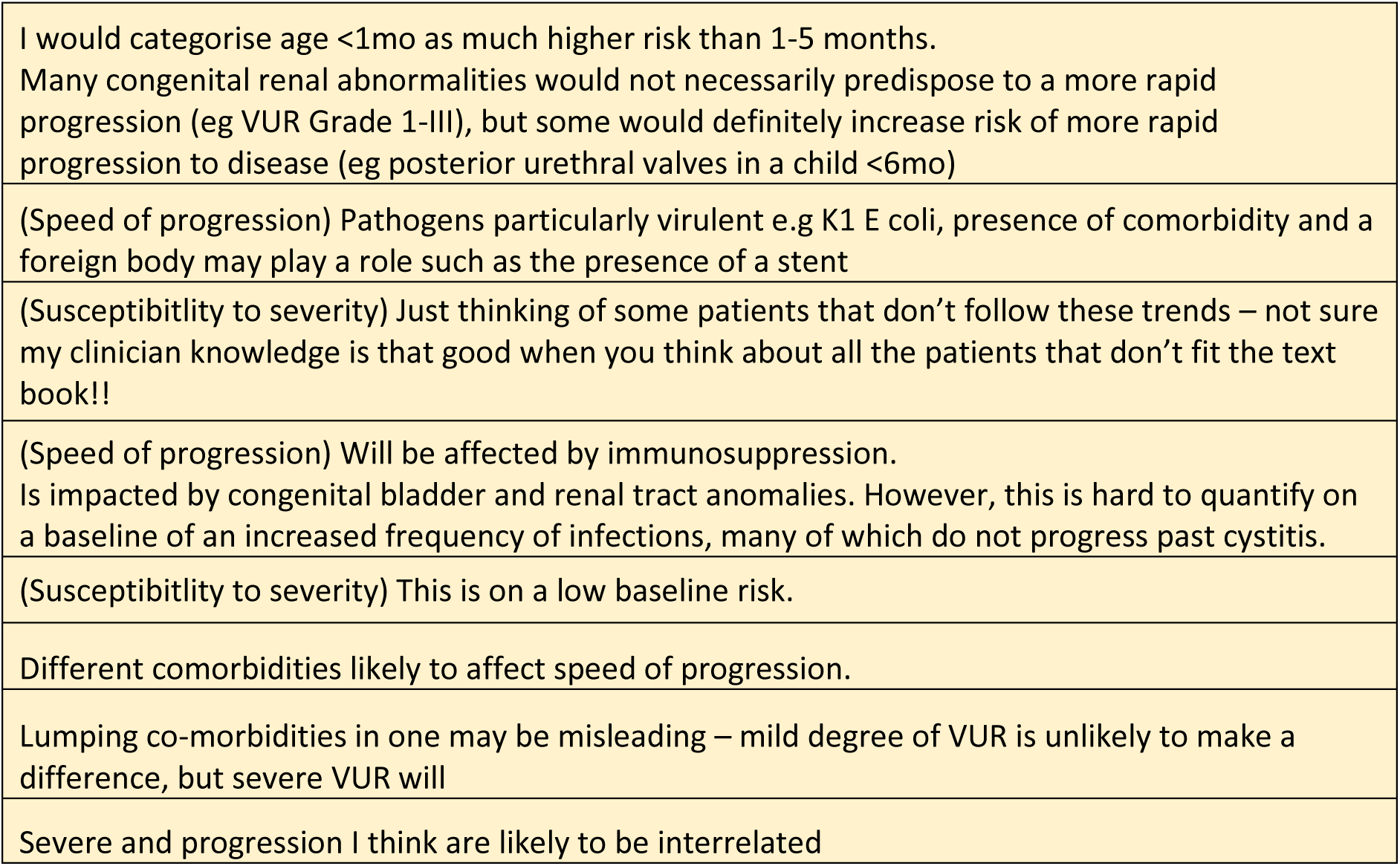

### Q3. Causative pathogen for UTI

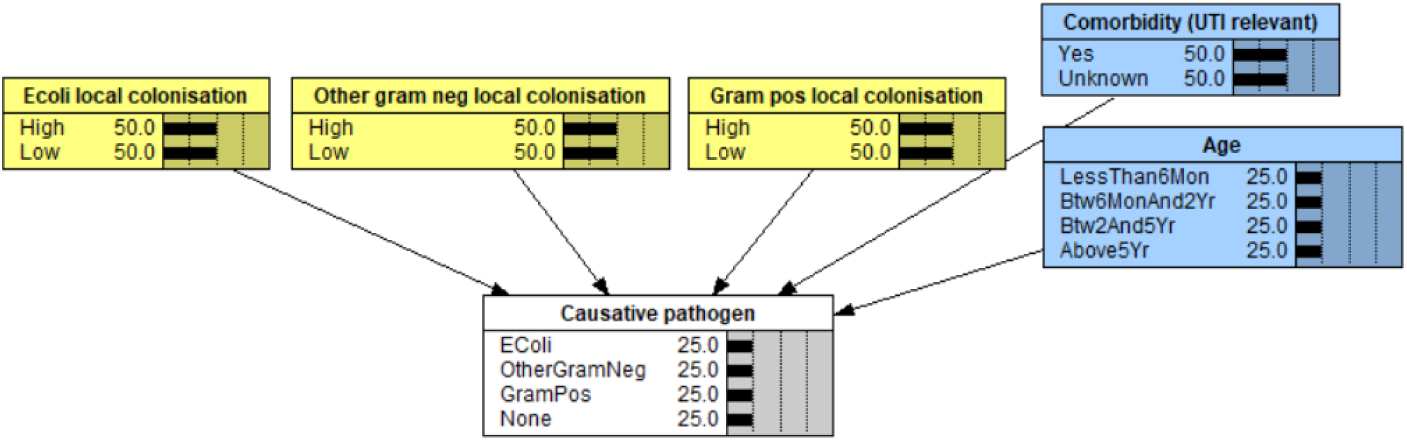

3a. Consider the PEA cohort, we enrolled children who presented to the Emergency Department (ED) at Perth Children’s Hospital and were managed for presumed UTI (with an antibiotic prescription in the ED and a urine sample sent for laboratory investigation). These patients typically underwent urine dipstick in the ED. What do you estimate <u>the probability</u> (min, max, best guess) of true UTI in this cohort (prior to seeing the laboratory culture result)? Please note that the “min/max” should be plausible lower or upper values, not the extreme recordable value. Mean and standard deviation based on survey responses from 8 experts.

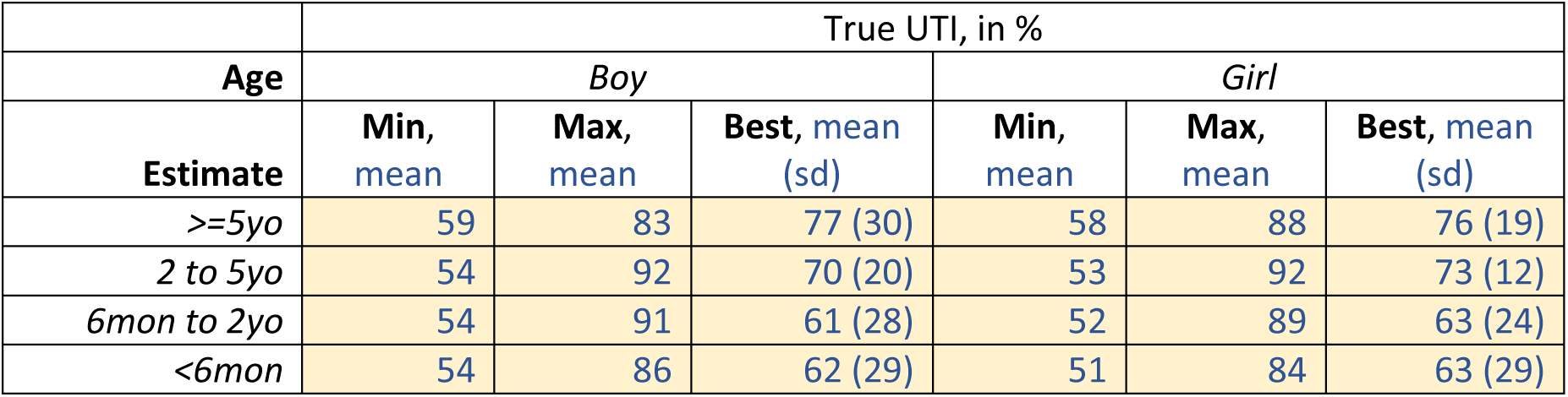

|  | True UTI, in % |  |  |  |  |  |
| --- | --- | --- | --- | --- | --- | --- |
| Age | Boy |  |  | Girl |  |  |
| Estimate | Min, mean | Max, mean | Best, mean (sd) | Min, mean | Max, mean | Best, mean (sd) |
| >=5yo | 59 | 83 | 77 (30) | 58 | 88 | 76 (19) |
| 2 to 5yo | 54 | 92 | 70 (20) | 53 | 92 | 73 (12) |
| 6mon to 2yo | 54 | 91 | 61 (28) | 52 | 89 | 63 (24) |
| <6mon | 54 | 86 | 62 (29) | 51 | 84 | 63 (29) |

Collated comments

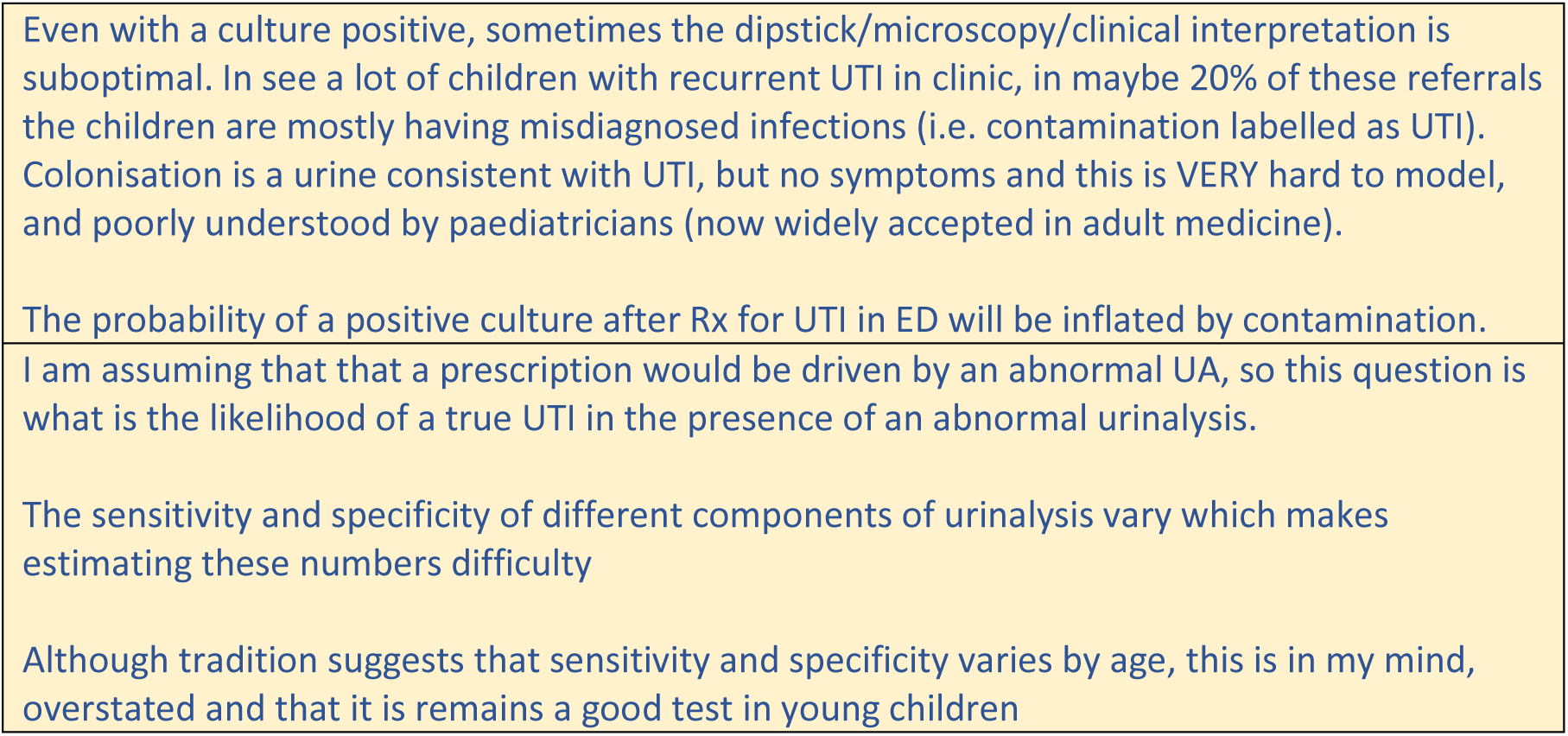

3b. In the case of an otherwise <u>healthy child</u> with colonisation of the perineum/ external genitalia by all the following three groups of organisms: E.coli, other gram negatives, and gram positives. Note, we refer to gram positives that can potentially cause UTI, such as Enterococcus, rather than gram positives like Staph epidermidis which are unlikely cause UTI. Mean and standard deviation based on survey responses from 8 experts.

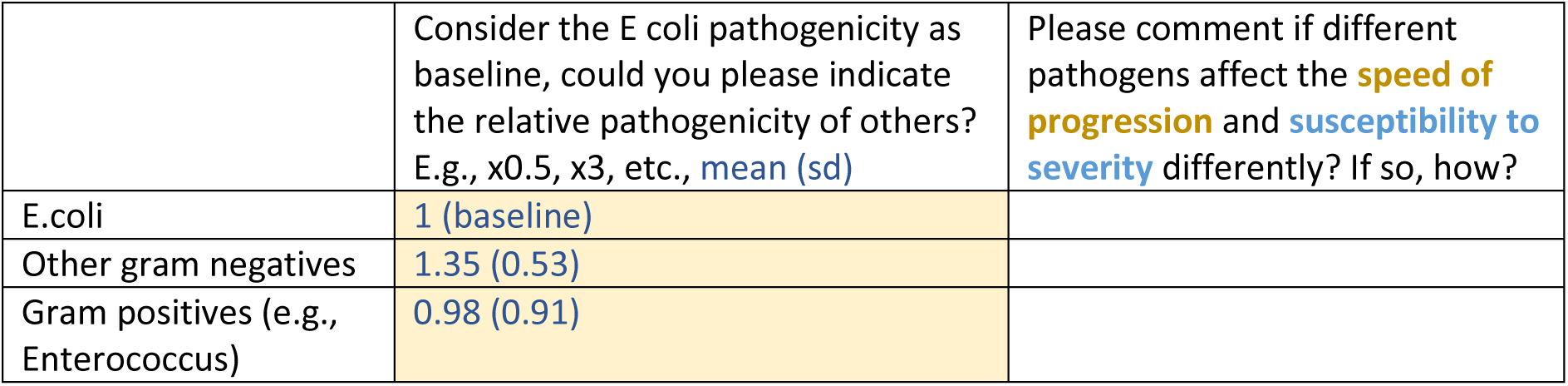

Collated comments

|  |
| --- |
| GramPos: Might be less than gram negatives, but do not have a good feel for this. |
| OtherGramNeg: Variable speed and severity (likely potentially lower, dependent on bug e.g., Kleb similar but Serratia lower) |
| GramPos: Low speed and severity |
| OtherGramNeg: Pseudomonas 2, Resistant gram negatives e.g ESBL 3; Particularly in a younger child, Given less likely to have early effective treatment and more challenging to treat |
| GramPos: Less severe and progressive |
| OtherGramNeg: Potentially more rapid spread with other gram negatives in the context of underlying renal anomalies and particularly with those with a stent in situ (biofilm formation, eg with organisms such as Pseudomonas aeruginosa). |
| GramPos: Generally less likely to cause disseminated infection. |
| E coli more frequently colonised, but the pathogenicity of this compared with other Enterobacteriaceae (e.g. Klebsiella, Enterobacter) is the same. Enterococci are the weeds of the urinary tract – in regard to pathogenicity, speed and severity |

### Q4. Impact of exiting antibiotic use

Comment: From only one expert.

|  |
| --- |
| Please see attached for a published list of antibiotics with their proposed “spectrum score”. Thus, I’d suggest you compare your below two groupings to ensure that all those in the “narrow” group have spectrum scores lower than all those in the “broader” group. Based on this you may need to move a few antibiotics between the two groups to ensure they are grouped appropriately (eg |

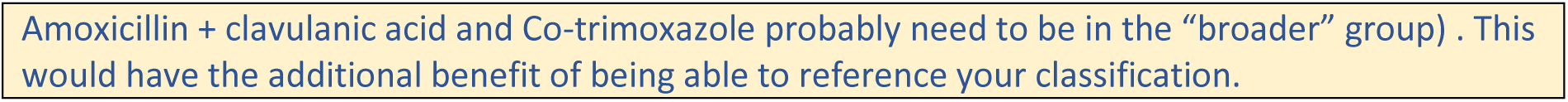

Under the following scenarios, please provide your min, max, and best guess estimates for each question. Please note that the “min/max” should be plausible lower or upper values, not the extreme recordable value. (Please feel free to refer to your experience of treating UTI in adults.) Response received from only one expert.

Consider a UTI caused by E.coli

|  | Probability of positive culture of E.coli |  |  |
| --- | --- | --- | --- |
| Estimate | Min | Max | Best guess |
| *Not on abx* | 50 | 100 | 80 |
| *On narrow abx* | 20 | 50 | 30 |
| *On broader abx* | 10 | 30 | 20 |
| *Pls feel free to add more groups* |  |  |  |

Consider a UTI caused by other gram negative bacteria

|  | Probability of positive culture of other gram neg |  |  |
| --- | --- | --- | --- |
| Estimate | Min | Max | Best guess |
| *Not on abx* | 50 | 100 | 80 |
| *On narrow abx* | 20 | 50 | 30 |
| *On broader abx* | 10 | 30 | 20 |

Consider a UTI caused by gram positive bacteria (e.g., Enterococcus)

|  | Probability of positive culture of gram pos |  |  |
| --- | --- | --- | --- |
| Estimate | Min | Max | Best guess |
| *Not on abx* | 50 | 100 | 80 |
| *On narrow abx* | 20 | 50 | 30 |
| *On broader abx* | 10 | 30 | 20 |

Comment:

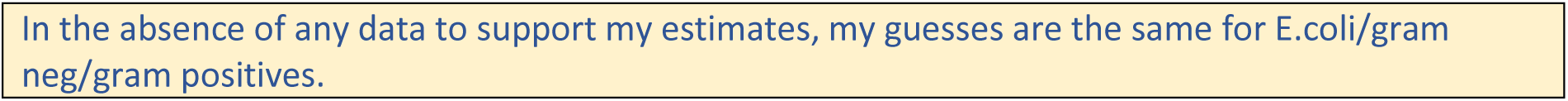

## Footnotes

1 The specified antibiotic was classified as narrow (<=3) or broader (>3) according to published Antibiotic Spectrum Index (22). Narrow: Amoxicilin, Trimethoprim, Benzylpenicillin, Cefalexin, Cefazolin, Erythromicin. Broad: Amoxicillin + Clavulanic acid, Trimethoprim + Sulfamethoxazole, Co-trimoxazole, Amikacin, Cefepime, Cefotaxime, Ceftazidime, Ceftriaxone, Ciprofloxacin, Colistin, Ertapenem, Gentamicin, Meropenem, Moxifloxacin, Nitrofurantoin, Norfloxacin, Piperacillin + Tazobactam, Tobramycin, Vancomycin

